# SARS-CoV-2 infection and post-acute risk of non-Covid-19 infectious disease hospitalizations: a nationwide cohort study of Danish adults aged ≥50 years

**DOI:** 10.1101/2023.04.03.23288102

**Authors:** Niklas Worm Andersson, Emilia Myrup Thiesson, Ria Lassaunière, Jørgen Vinsløv Hansen, Anders Hviid

## Abstract

Reports suggest that the potential long-lasting health consequences of SARS-CoV-2 infection may involve persistent dysregulation of some immune populations, but the potential clinical implications are unknown. In a nationwide cohort of 2,430,694 50+-year-olds, we compared the rates of non-Covid-19 infectious disease inpatient hospitalizations (of ≥5 hours) following the acute phase of SARS-CoV-2 infection in 930,071 individuals with rates among SARS-CoV-2 uninfected from 1 January 2021 to 10 December 2022. The post-acute phase of SARS-CoV-2 infection was associated with an incidence rate ratio of 0.90 (95% confidence interval 0.88-0.92) for any infectious disease hospitalization. Findings were similar for upper- (1.08, 0.97-1.20), lower respiratory tract (0.90, 0.87-0.93), influenza (1.04, 0.94-1.15), gastrointestinal (1.28, 0.78-2.09), skin (0.98, 0.93-1.03), urinary tract (1.01, 0.96-1.08), certain invasive bacterial (0.96, 0.91-0.1.01), and other (0.96, 0.92-1.00) infectious disease hospitalizations and in subgroups. Our study does not support an increased susceptibility to non-Covid-19 infectious disease hospitalization following SARS-CoV-2 infection.

## INTRODUCTION

The introduction of Covid-19 into human society has had a profound impact on global health, with hundreds of millions of people infected worldwide.^1^ A considerable number of individuals infected with SARS-CoV-2 continue to suffer from persisting symptoms following the acute phase of their illness.^2–6^ These long-term symptoms have been reported to be multifaceted including both physiological and psychological manifestations such as debilitating fatigue, dyspnea, chest pain, headaches, and cognitive dysfunction.^6–8^ Collectively, known as Post-Covid-19 condition or long Covid and shown to have significant impact on quality of life.^5–9^

While our knowledge of the long-term health consequences of Covid-19 continues to advance, the full range of implications is not yet fully understood. In addition, SARS-CoV-2 infection may have a prolonged effect on both the innate and adaptive immune system.^10–19^ The affected immune compartments and duration of dysregulation after Covid-19 include reduced absolute numbers of CD4+ T cells, CD8+ T cells and NK cells for up to 2.5-3 months^10–12,14^, increased activation of CD4+ and CD8+ T cells for up to 12 months^12–15^, and reduced plasmacytoid dendritic cells for up to 7 months^16^. Moreover, post-acute Covid-19 immune perturbations seem most pronounced up to 3 months after SARS-CoV-2 infection^10^, associated with older age and severe Covid-19.^12,13^ Nonetheless, the clinical consequence of these potential immunological dysregulations in terms of risk of other severe non-Covid-19 infectious diseases remains unclear.

We leveraged the comprehensive Danish test- and surveillance system for Covid-19 together with nationwide healthcare- and demography registers to investigate the associated risk of non-Covid-19 infectious disease hospitalizations after the acute phase of SARS-CoV-2 infection in a large national cohort of adults aged ≥50 years.

## RESULTS

Among the 2,430,694 included individuals (mean age 66.8 years, standard deviation 11.3 years), 930,071 individuals were infected with SARS-CoV-2 (ascertained by first positive PCR test) during the study period from 1 January 2021 to 10 December 2022 (Table 1 and Supplementary Figure S1). Follow-up for the entire study cohort totaled 4,519,913 person-years. Slightly more than half (51.8%) of the population were females. Demography-, comorbidity-, and vaccination status characteristics were similar between the entire study cohort and those acquiring a SARS-CoV-2 infection. We identified a total of 78,555 infectious disease hospitalizations of any type during follow-up (Figure 1 and Figure 2). Examining the individual secondary outcomes, lower respiratory tract infections were the most common cause of hospitalization, followed by the other types of infection-, urinary tract infection-, and certain invasive bacterial infection categories.

**Table 1.**
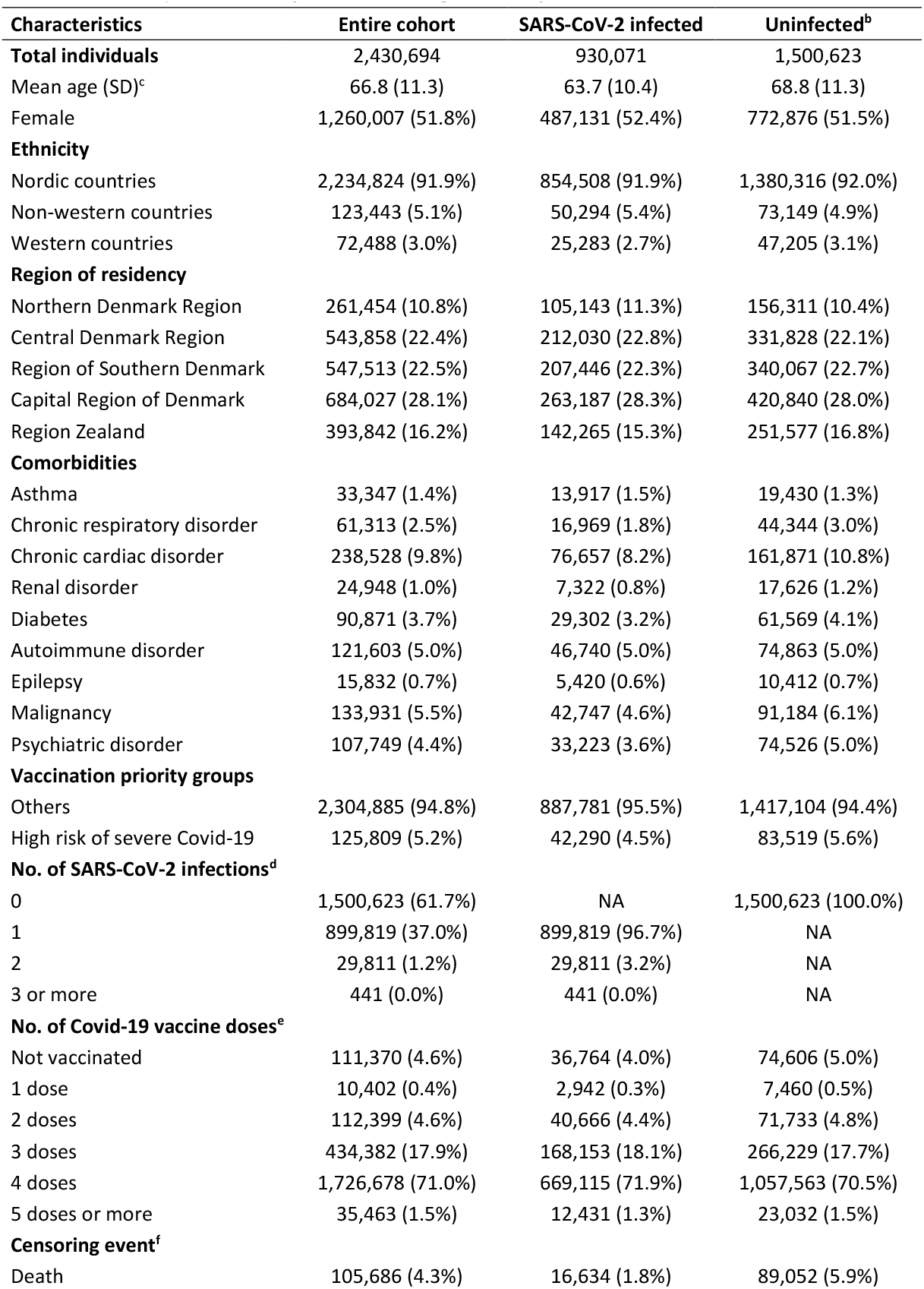
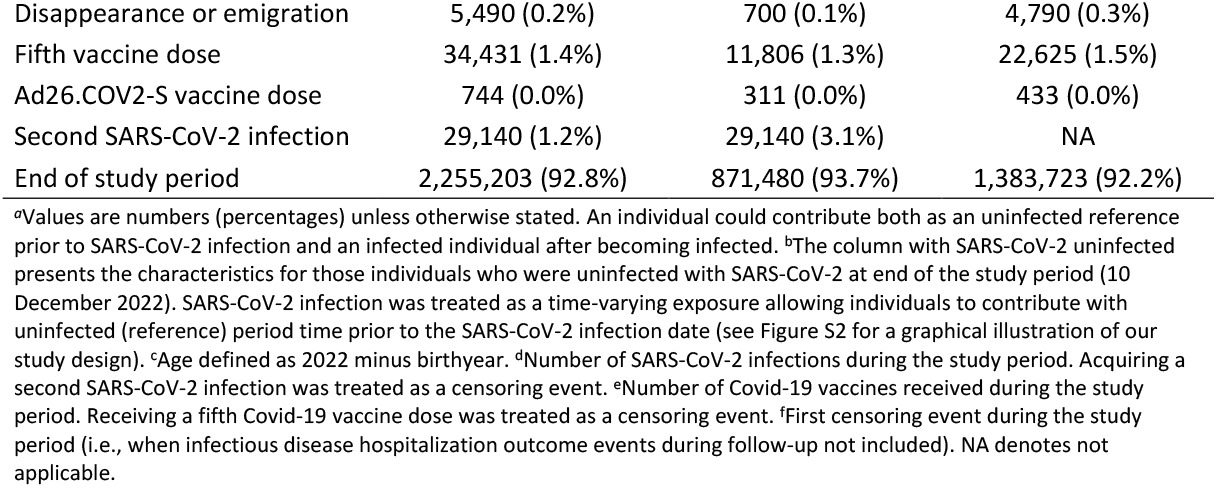
Characteristics of the entire cohort, SARS-CoV-2 infected, and uninfected (at 10 December 2022) Danish 50+ year-olds during 1 January 2021 to 10 December 2022.^*a*^.

**Figure 1.**
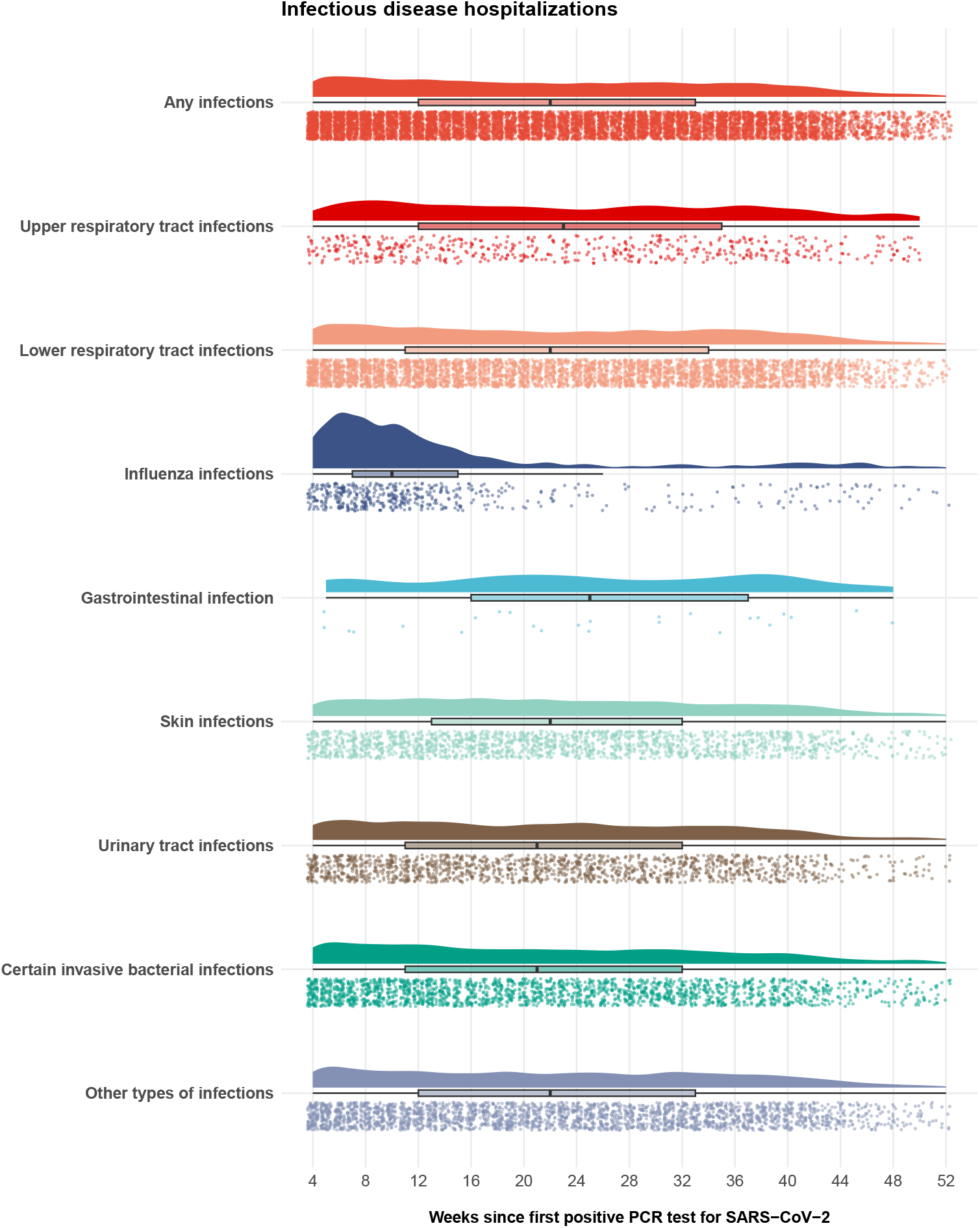
Distribution of infectious disease hospitalizations by weeks during main risk period from day 29+ following first SARS-CoV-2 infection. The distribution of the primary and secondary non-Covid-19 infectious disease hospitalization outcomes between week 4 (day 29) through week 52 (day 365) after the SARS-CoV-2 infection date (day 0) are presented by half-density (raincloud) plots, box plots, and jitter points.

**Figure 2.**
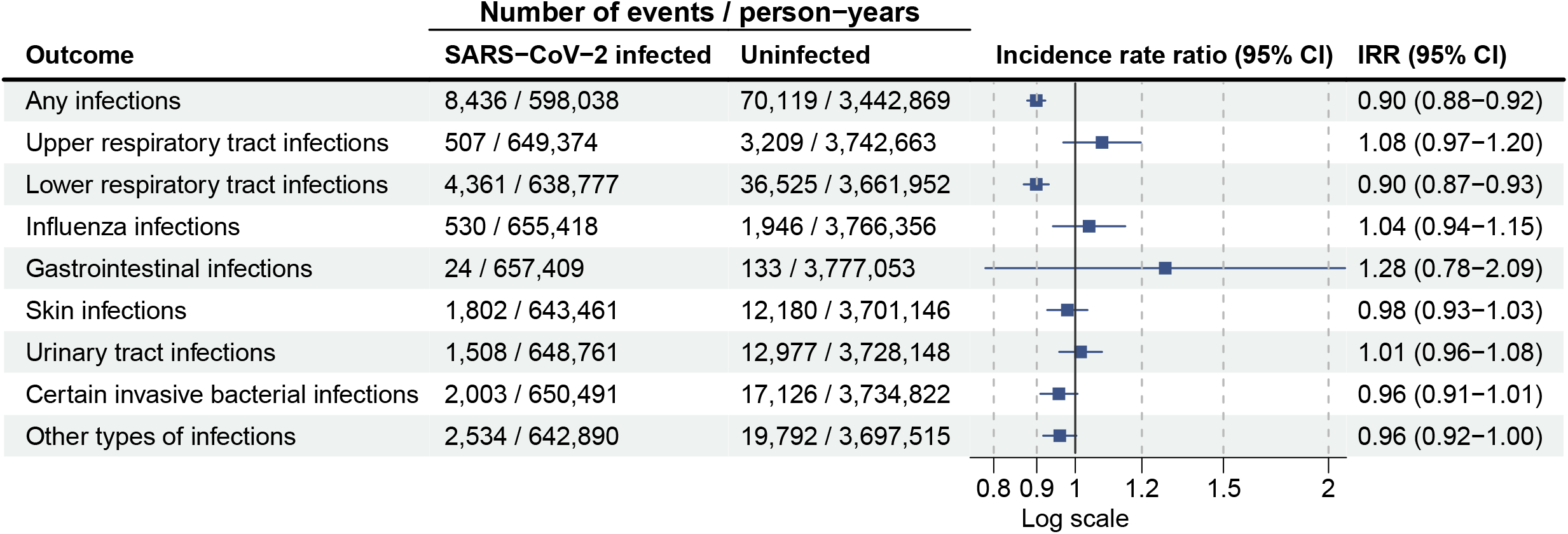
Risk of infectious disease hospitalization after +29 days since first SARS-CoV-2 infection in Danish 50+ year-olds and uninfected reference period during 1 January 2021 to 10 December 2022. CI denotes confidence interval and IRR incidence rate ratio.

Comparing the main risk period of day 29+ following SARS-CoV-2 infection with the SARS-CoV-2 uninfected reference period for any infectious disease hospitalization, the incidence rate ratio (IRR) was 0.90 (95% confidence interval [CI] 0.88-0.92) (Figure 2). Likewise, no increased risks were observed for secondary outcomes when individually assessed. The IRR was 1.08 (0.97-1.20) for upper respiratory tract, 0.90 (0.87-0.93) for lower respiratory tract, 1.04 (0.94-1.15) for influenza, 1.28 (0.78-2.09) for gastrointestinal, 0.98 (0.93-1.03) for skin, 1.01 (0.96-1.08) for urinary tract, 0.96 (0.91-1.01) for certain invasive bacterial, and 0.96 (0.92-1.00) other types of infectious disease hospitalization. No major differences in the risks were identified when stratifying the cohort according to sex and age subgroups (Table S1). For the age group consisting of individuals aged ≥80 years, we observed an IRR of 0.96 (0.92-1.00) for any infectious disease hospitalization, but the IRR was 1.22 (1.02-1.45) for influenza, 1.16 (1.03-1.30) for skin, 1.17 (1.07-1.27) for urinary tract, and 1.13 (1.04-1.23) for certain invasive bacterial infectious disease hospitalization.

Examining the risk of infectious disease hospitalizations according to vaccination status at time of SARS-CoV-2 infection was not suggestive of consistent differential risks (Figure 3). The IRR for any infectious disease hospitalization was 0.95 (0.88-1.02) for unvaccinated, 0.93 (0.87-0.99) for primary course vaccinated, and 0.88 (0.86-0.91) for booster vaccinated at time of SARS-CoV-2 infection as compared with SARS-CoV-2 uninfected with similar vaccination status. Comparing the risk of any infectious disease hospitalization according to vaccination status did not change across sex and age subgroups (Table S2). For the individual infectious disease hospitalizations, however, we observed tendencies toward significant associations in some of the sex and age subgroup analyses among unvaccinated and primary course vaccinated individuals; however, these estimates were based on lower numbers of cases among SARS-CoV-2 infected. Among booster vaccinated, no associated risks were seen for SARS-CoV-2 infected, including among those individuals aged ≥80 years.

**Figure 3.**
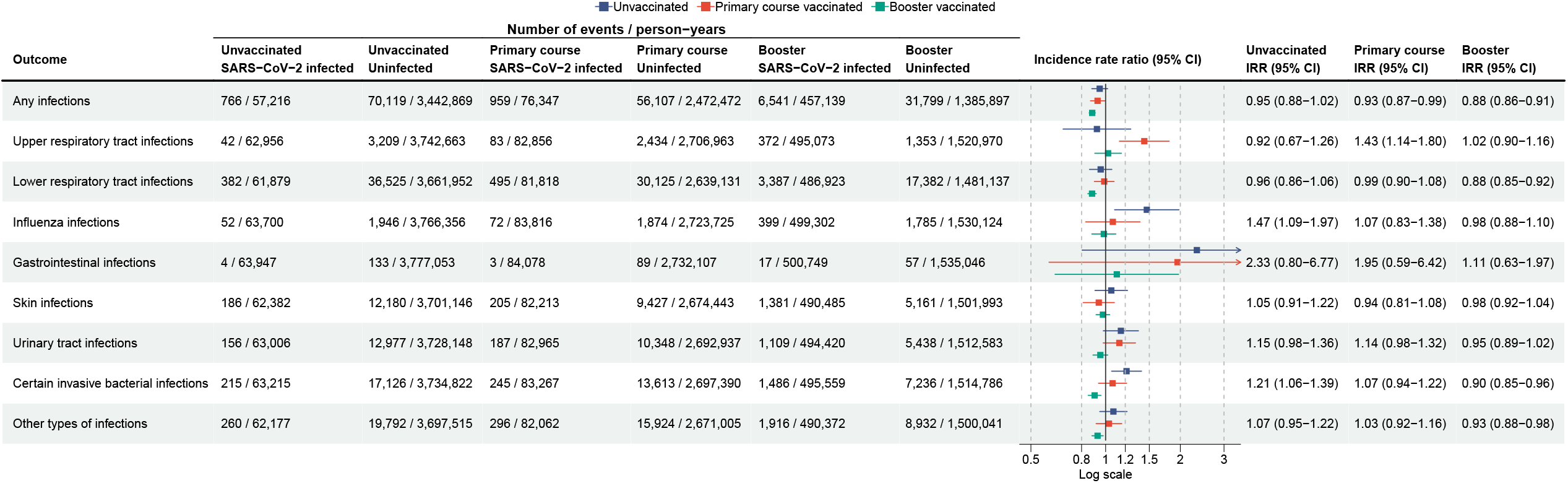
Risk of infectious disease hospitalization after 29+ days since first SARS-CoV-2 infection in Danish 50+ year-olds and uninfected reference period during 1 January 2021 to 10 December 2022 according to Covid-19 vaccination status at time of SARS-CoV-2 infection.^**a**^. ^a^Unvaccinated (zero doses), primary course vaccinated (two doses, i.e., time between first and second dose was left out), and booster (three or four doses) denotes the number Covid-19 vaccines received at time of the first SARS-CoV-2 infection. SARS-CoV-2 infected were compared with uninfected (reference) individuals with similar vaccination status. A first SARS-CoV-2 infection after any dose was treated as a censoring event for the unvaccinated comparison as was a first SARS-CoV-2 infection after the third dose for the primary course vaccinated comparison (see Figure S3). CI denotes confidence interval and IRR incidence rate ratio.

Splitting the post-acute period into day 29-180 and >day 180 risk windows since the SARS-CoV-2 infection date, showed similar results to those of the primary analysis; the IRR for any infection was 0.93 (0.90-0.96) for day 29-180 and 0.86 (0.83-0.89) for >180 days (Table 2). Likewise, estimates were largely unchanged in sensitivity analyses deferring the start of the main risk period to day 90 since SARS-CoV-2 infection (IRR 0.86, 0.84-0.89 for any infectious disease hospitalization, Table S3) and applying a pre-risk period of -7 days before the first SARS-CoV-2 infection date (IRR 0.93, 0.91-0.95 for any infectious disease hospitalization, Table S4).

**Table 2.** Risk of infectious disease hospitalization after 29-180 days and >180 days since first SARS-CoV-2 infection in Danish 50+ year-olds and reference period from 1 January 2021 to 10 December 2022.

| Outcome | Number of events / person-years |  |  | Incidence rate ratio (95% CI) |  |
| --- | --- | --- | --- | --- | --- |
|  | SARS-CoV-2 infected |  | Uninfected | 29-180 days | >180 days |
|  | 29-180 days | >180 days |  |  |  |
| Any infections | 4,828 / 328,561 | 3,608 / 269,478 | 70,119 / 3,442,869 | 0.93 (0.90-0.96) | 0.86 (0.83-0.89) |
| Upper respiratory tract infections | 274 / 356,566 | 233 / 292,808 | 3,209 / 3,742,663 | 1.02 (0.89-1.17) | 1.15 (0.99-1.34) |
| Lower respiratory tract infections | 2,474 / 350,581 | 1,887 / 288,195 | 36,525 / 3,661,952 | 0.94 (0.90-0.99) | 0.84 (0.80-0.89) |
| Influenza infections | 453 / 359,792 | 77 / 295,625 | 1,946 / 3,766,356 | 1.01 (0.91-1.12) | 1.24 (0.97-1.58) |
| Gastrointestinal infections | 12 / 360,880 | 12 / 296,529 | 133 / 3,777,053 | 1.11 (0.59-2.12) | 1.53 (0.77-3.01) |
| Skin infections | 1,041 / 353,378 | 761 / 290,082 | 12,180 / 3,701,146 | 1.02 (0.95-1.09) | 0.92 (0.85-1.00) |
| Urinary tract infections | 896 / 356,174 | 612 / 292,586 | 12,977 / 3,728,148 | 1.06 (0.99-1.14) | 0.95 (0.87-1.04) |
| Certain invasive bacterial infections | 1,192 / 357,068 | 811 / 293,424 | 17,126 / 3,734,822 | 1.03 (0.96-1.09) | 0.87 (0.80-0.93) |
| Other types of infections | 1,433 / 353,005 | 1,101 / 289,885 | 19,792 / 3,697,515 | 1.00 (0.94-1.06) | 0.91 (0.85-0.97) |

CI denotes confidence interval.

To inform previous findings^20^, we observed no increased risk of infectious disease hospitalization for tonsillitis in the 29-180- and >180-day periods following SARS-CoV-2 infection, nor when stratified by sex or age subgroups (Table S5). As expected, the rates of the infectious disease hospitalization outcomes following day 90+ of Covid-19 hospitalization were universally increased compared with SARS-CoV-2 uninfected reference period rates (IRR 2.28, 2.08-2.49 for any infectious disease hospitalization, Table S6).

## DISCUSSION

We estimated the subsequent risk of hospitalization with non-Covid-19 infectious diseases following recovery from SARS-CoV-2 infection in a nationwide cohort of Danish adults aged ≥50 years from 1 January 2021 to 10 December 2022. Our results do not support that SARS-CoV-2 infection increases the susceptibility to non-Covid-19 infectious diseases in this population.

Given the observed persistent dysregulations of innate and adaptive immune populations in SARS-CoV-2 infected compared to healthy controls,^10,15–18^ the theoretical concern is not unjustified. The reduced levels of dendritic cells may imply an impaired ability to initiate immune responses against new pathogens, while prolonged lymphopenia may increase risk for hospitalization with infectious diseases.^21^ However, studies of the immunological sequelae observed after an SARS-CoV-2 infection are limited to peripheral immune cells in unvaccinated individuals, with immune dysregulation primarily occurring after severe Covid-19.^10,12,14–19^ It remains unclear if post-acute Covid-19 modulation extends to tissue resident immune cells, that are present at the site of new infections, or vaccinated individuals with breakthrough SARS-CoV-2 infections. Data to evaluate how post-acute Covid-19 immunological perturbations (known and unknown) in unvaccinated and vaccinated individuals may translate into clinical consequences, however, are not well-established.

Compared with SARS-CoV-2-test-negative controls, a Danish population sample of 9893 unvaccinated, non-hospitalized SARS-CoV-2 positive cases from 27 February 2020 to 31 May 2020, reported no increased risk of any medication use or diagnoses related to infectious disease from 14 to 180 days after the SARS-CoV-2 test in a hypothesis-free screening analysis (screened on the fifth level of the Anatomical Therapeutical Chemical classification for drug substances and second level of ICD-10 codes for diagnoses).^22^ One retrospective Israeli cohort study examined the risk of conjunctivitis, streptococcal tonsillitis, pneumonia, Epstein-Barr virus, and herpes virus infections (that is, not restricted to inpatient hospitalization) among other long Covid-19 outcomes in an unvaccinated SARS-CoV-2 infected (a total of 170,280 infected during Index or Alpha subvariant predominance periods) vs an uninfected population, consisting primarily of younger individuals (median age 24 years, interquartile range 13-42 years; 10,835 [6.4%] were aged >60 years).^20^ While their main results observed a hazard ratio of 1.18 (95% CI, 1.09-1.28) and 1.12 (1.05-1.20) for streptococcal tonsillitis for the assessed intervals of day 30-180 and day 180-360 since SARS-CoV-2 infection, respectively, no other increased risks of other infections were found. In individuals aged 41-60 and >60 years, no increased infection risks were observed (except for tonsillitis among 41-60-year-olds from 180-360 days following infection).^20^ Thus, with the exception of a modest increase in streptococcal tonsillitis, these studies of unvaccinated individuals do not support an overall increased risk of microbial infectious diseases during the period of post-Covid-19 lymphopenia and other immune perturbations.

Rather, since the immune memory responses against non-SARS-CoV-2 are maintained following a SARS-CoV-2 infection^14,23^, these responses may contribute to sustained protection against infectious diseases^24^; unlike the measles virus-associated loss of adaptive immune memory, that associates with a long-lasting effect on the risk of acquiring infections with other pathogens.^25–30^ None of the aforementioned studies included Covid-19 vaccinated individuals.^10–20,22^ We found no evidence of an increased risk of non-Covid-19 infectious disease hospitalization associated with SARS-CoV-2 infection in individuals having received one or two Covid-19 vaccine booster doses. As such, our study provides a pertinent evaluation that may inform booster vaccinations.

The inherent methodological boundaries of observational data should be reflected in the clinical interpretation of our findings. Particularly, the possibility that behavioral and environmental differences between SARS-CoV-2 infected and uninfected individuals may exist. One possible factor could be differences in risk behavior (such as being less precautious and more socially interactive) of SARS-CoV-2 infected individuals relative to uninfected at that time, leading to differences in the susceptibility of acquiring other transmittable diseases. Additionally, patterns in healthcare seeking behavior may also be different between SARS-CoV-2 infected and uninfected. Similarly, given the increased public as well as clinical awareness of symptoms related to SARS-CoV-2 infection, risk estimates of other infectious diseases with similar clinical manifestations to those of SARS-CoV-2, such as other upper respiratory tract infections, would likely be mostly influenced by potential differences in healthcare seeking patterns. In the context of examining the risk of severe infections these potential biases from differences in risk- and healthcare seeking behavior are mitigated, but would otherwise most likely tend to skew estimates toward increased risks for SARS-CoV-2 infected. Still, our inpatient hospitalization definition of hospital contacts of ≥5 hours was most likely subject to some outcome misclassification (i.e., also capturing some less severe outcome events). Our exposure definition relied on a registered positive PCR test for SARS-CoV-2, and hence, holds potential for exposed being misclassified as unexposed; however, extensive *free-of-charge* national SARS-CoV-2 testing strategies were implemented in Denmark (e.g., the number of SARS-CoV-2 PCR tests performed per capita far exceeds most other countries^31^) during the Covid-19 pandemic. Moreover, given the observational nature of our study residual confounding cannot be ruled out to the extent that confounders not adjusted for were differently distributed between compared periods. Our study results were primarily null findings; however, we did observe tendencies toward some signals among individuals aged ≥80 years as well as for those hospitalized for Covid-19, that is, subgroup populations with general greater background prevalence of comorbidity and frailty that may be difficult to adequately capture with our utilized observational design. As such, while our inclusion of individuals of these older ages increases the generalizability of our results, the degree of residual confounding was likely greater for this specific subgroup. Similarly, as expected, the risk of hospitalization for other infectious diseases was universally increased among Covid-19 hospitalized compared with uninfected. Lastly, our secondary outcome analyses for individuals infected with SARS-CoV-2 infected among unvaccinated and after receiving a primary vaccination course-only was limited by lower statistical precision, but we observed no consistent risk patterns across outcomes or subgroups. Notably, comparing the risk in booster vaccinated only, an analysis where the internal validity was maximized, detected no associations for SARS-CoV-2 infected in any outcome or subgroup analysis.

Owing to the nationwide coverage of the longitudinal individual-level Danish healthcare register data, selection bias is minimized and allows for a high degree of generalizability to similar populations. As we studied the risk of infectious disease hospitalization among Danish adults aged ≥50 years, our results may have less applicability in evaluations of associated risks in younger or demographically different populations. Likewise, the relevancy of our results to other infectious diseases or clinical scenarios not studied, such as individuals having had multiple SARS-CoV-2 infections, is unknown.

In conclusion, in this nationwide cohort study from 1 January 2021 to 10 December 2022 of Danish adults aged ≥50 years, SARS-CoV-2 infection was not associated with an increased long-term susceptibility to inpatient hospitalization with non-Covid-19 infectious diseases.

## ONLINE METHODS

### Setting and study population

We cross-linked several Danish test- and surveillance system for Covid-19 with healthcare- and demography registers, all with nationwide coverage, using the unique identifier assigned to all residents in Denmark at either birth or immigration.^32–35^ This allowed us to obtain data on SARS-CoV-2 infection, hospitalization with physician-assigned diagnoses (according to the International Classification of Disease, revision 10), Covid-19 vaccination status, and other covariates on an individual level. Supplementary Tables S7 and S8 provide details on the utilized data sources and variable definitions. According to Danish law, register-based research in Denmark is exempt from ethics committee approval.

During the study period 1 January 2021 to 10 December 2022, we constructed a cohort representative of the general Danish population born in 1972 or earlier (i.e., equal to turning at least 50 years in 2022). Additionally, to be included in our study cohort, we required individuals to have Danish residency at baseline and not having been infected with SARS-CoV-2 prior to study entry. SARS-CoV-2 infection was defined as a registered positive PCR test for SARS-CoV-2 and was used to create a time-varying exposure. We designated the first 28 days after the infection day (day 0) as the acute phase of individuals’ first SARS-CoV-2 infection while our main risk period of interest was day 29 following infection and onwards. Time as SARS-CoV-2 uninfected constituted as the reference period (see Figure S2 for a graphical scheme of our study design).

### Outcomes

Study outcomes in the form of incident events of infectious disease hospitalization were defined by primary or secondary diagnoses of infectious disease (adapted and modified from previous work^36^; Table S8) assigned during hospital contacts of ≥5 hours. The main outcome was any infectious disease hospitalization, while secondary outcomes were constructed by subclassifying the diagnoses into upper respiratory tract, lower respiratory tract (influenza not included), influenza, gastrointestinal, skin, urinary tract, certain invasive bacterial (i.e., sepsis, meningitis, endocarditis, and osteomyelitis), and other types of infectious diseases. We used the date of admission as the date of the event and studied each individual outcome separately. We excluded individuals who had a recent history of the respective outcome during a washout period from 1 January 2018 to 31 December 2020.

### Statistical analysis

We followed individuals from study entry (i.e., 1 January 2021 or turning age 50 years, whichever occurred last) until first outcome event, second positive PCR test for SARS-CoV-2, emigration, death, receipt of a fifth vaccine dose (as not rolled out to the general Danish population during the study period), or 10 December 2022, whichever occurred first. We also right-censored individuals upon the day of receiving an Ad26.COV2-S Covid-19 vaccine as these were few within our study cohort and/or not recommended for the studied population. Infected individuals could contribute with person-time during both the SARS-CoV-2 uninfected (reference), acute SARS-CoV-2 infection phase (1 to 28 days following first SARS-CoV-2 infection date; estimates not reported), and main risk (≥29 days following first SARS-CoV-2 infection; i.e., post-acute phase) period. Comparing the outcome rates during the main risk (≥29 days since infection) and uninfected reference period, we estimated adjusted incidence rate ratios (IRRs) with corresponding 95% confidence intervals (CI) from a log-linear Poisson regression with the logarithm of the follow-up time as the offset. The model was adjusted for sex, age (in 5-year bins), ethnicity (Nordic, Western, non-Western), region of residence (5 levels), considered at high risk of severe Covid-19 (binary), calendar time (biweekly), vaccination status (5 levels), and number of comorbidities (defined as asthma, other chronic respiratory disorders, chronic cardiac disorders, renal disorders, diabetes, autoimmune-related disorders, epilepsy, malignancies, and psychiatric disorders; summed to: 0, 1, ≥2 comorbidities). Age, calendar time, and vaccination status was treated as time-varying covariates, the others were ascertained at baseline.

In secondary analyses, we examined the associations by sex and age (50-64, 65-79, and ≥80 years) subgroups, by Covid-19 vaccination status at time of SARS-CoV-2 infection (separate analyses were made for unvaccinated, primary course vaccinated [received two vaccine doses], and booster vaccinated [received one or two booster doses]) and by splitting the main risk period at day 180 (i.e., day 29-180+ and >180+) after the SARS-CoV-2 infection date. For the analyses of Covid-19 vaccination status, individuals were excluded or right-censored if acquiring SARS-CoV-2 infection prior to or after the particular vaccination status-time analyzed, respectively (see Figure S3 for a graphical illustration of these analyses). In sensitivity analyses, we deferred the time of start of follow-up to day 90 since the day of SARS-CoV-2 infection and applied a pre-risk period of -7 days to the date of first SARS-CoV-2 infection. For the latter, this pre-risk period was omitted from the uninfected (reference) period (to assess for any influence by the uncertainty of the specific order of an outcome and exposure during this period [including day 0]). To inform and expand on previous study findings,^20^ we examined the risk of infectious disease hospitalization for tonsillitis (day 29-180+ and >180+ after SARS-CoV-2 infection) separately. Lastly, we compared the rates of the outcomes following Covid-19 hospitalization (≥90+ days from the admission date) with the uninfected outcome rates.

Statistical tests were 2-sided; associations were considered statistical significance if the 95% CI did not overlap with 1. SAS version 9.4 and R version 4.0.2 was used for data management, while statistical analysis was completed in R version 4.0.2.

## Data Availability

No additional data available. Owing to data privacy regulations in Denmark, the raw data cannot be shared.

## Transparency

The lead author (the manuscript’s guarantor) affirms that this manuscript is an honest, accurate, and transparent account of the study being reported; that no important aspects of the study have been omitted; and that any discrepancies from the study as planned (and, if relevant, registered) have been explained.

## Ethics

The analyses were performed as surveillance activities analyses as part of the advisory tasks of the governmental institution Statens Serum Institut (SSI) for the Danish Ministry of Health. SSI’s purpose is to monitor and fight the spread of disease in accordance with section 222 of the Danish Health Act. According to Danish law, national surveillance activities conducted by SSI do not require approval from an ethics committee. Both the Danish Governmental law firm and the compliance department of SSI have approved that the study is fully compliant with all legal, ethical, and IT-security requirements and there are no further approval procedures required for such studies.

