## Supplementary Figure S1 for "SARS-CoV-2 infection and post-acute risk of non-Covid-19 infectious disease hospitalizations: a nationwide cohort study of Danish adults aged ≥50 years"

#### Table of contents

Supplementary Figure S1. Distribution of first SARS-CoV-2 infected (ascertained by positive PCR tests) during the study period from 1 January 2021 to 10 December 2022, presented by density plot, box plot, and heat map. 2

Supplementary Table S1. Risk of infectious disease hospitalization after 29+ days since first SARS-CoV-2 infection in Danish 50+ year-olds and uninfected reference period during 1 January 2021 to 10 December 2022 by sex and age subgroups. 3

Supplementary Table S2. Risk of infectious disease hospitalization after 29+ days since first SARS-CoV-2 infection in Danish 50+ year-olds and uninfected reference period during 1 January 2021 to 10 December 2022 according to Covid-19 vaccination status at time of SARS-CoV-2 infection by sex and age subgroups.[a] 5

Supplementary Table S3. Sensitivity analysis of risk of infectious disease hospitalization where using an alternative main risk period of ≥90+ days since first SARS-CoV-2 infection. 10

Supplementary Table S4. Sensitivity analysis of risk of infectious disease hospitalization where using pre-risk period of -7 days prior to the first SARS-CoV-2 infection. 12

Supplementary Table S5. Risk of infectious disease hospitalization with tonsillitis after ≥29+ days since first SARS-CoV-2 infection during 1 January 2021 to 10 December 2022. **Fejl! Bogmærke er ikke defineret.**

Supplementary Table S6. Risk of infectious disease hospitalization after 90+ days since first Covid-19 hospitalization in Danish 50+ year-olds and uninfected reference period during 1 January 2021 to 10 December 2022.[a] 15

Supplementary Table S7. Eligibility criteria and exposure and covariates definitions. 17

Supplementary Table S8. Infectious disease hospitalization outcome definitions. 20

Supplementary Figure S2. Schematic figure of the study design. 22

Supplementary Figure S3. Schematic figure of the analyses comparing SARS-CoV-2 infected and uninfected reference according to vaccination status at time of infection. 23

Supplementary References 24

**Supplementary Figure S1. Distribution of first SARS-CoV-2 infected (ascertained by positive PCR tests) during the study period from 1 January 2021 to 10 December 2022, presented by density plot, box plot, and heat map.**

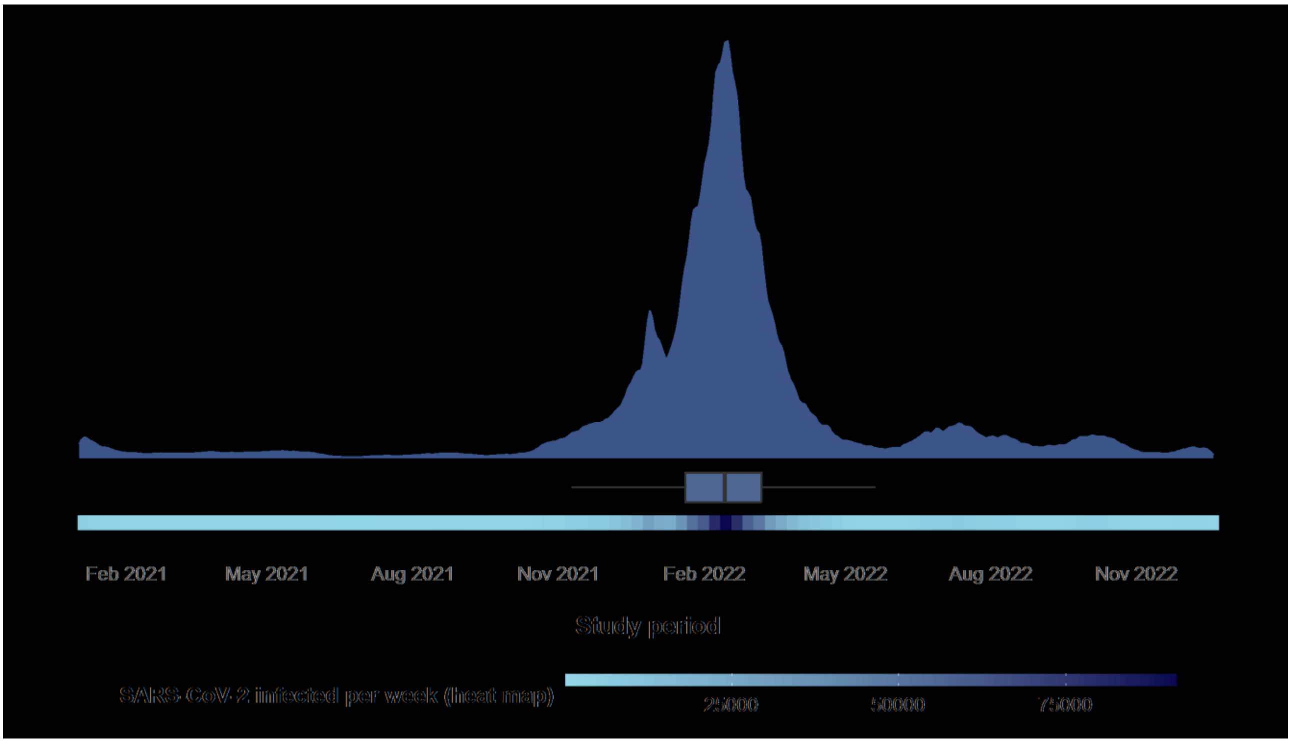

**Supplementary Table S1. Risk of infectious disease hospitalization after 29+ days since first SARS-CoV-2 infection in Danish 50+ year-olds and uninfected reference period during 1 January 2021 to 10 December 2022 by sex and age subgroups.**

| Outcome event / subgroup | Number of events / person-years | | Incidence rate ratio (95% CI) |
|---|---|---|---|
| | SARS-CoV-2 infected | Uninfected | |
| **Any infections** | | | |
| Female | 3,771 / 314,099 | 32,830 / 1,795,033 | 0.88 (0.85-0.91) |
| Male | 4,665 / 283,940 | 37,289 / 1,647,836 | 0.92 (0.89-0.95) |
| Aged 50 to 64 years | 2,568 / 379,743 | 12,690 / 1,606,402 | 0.86 (0.82-0.90) |
| Aged 65 to 79 years | 3,444 / 180,538 | 29,039 / 1,380,960 | 0.92 (0.89-0.96) |
| Aged 80 years or older | 2,424 / 37,758 | 28,390 / 455,507 | 0.96 (0.92-1.00) |
| **Upper respiratory tract infections** | | | |
| Female | 245 / 338,938 | 1,532 / 1,941,164 | 0.99 (0.85-1.15) |
| Male | 262 / 310,435 | 1,677 / 1,801,499 | 1.15 (0.99-1.33) |
| Aged 50 to 64 years | 292 / 402,365 | 1,164 / 1,701,058 | 1.13 (0.97-1.31) |
| Aged 65 to 79 years | 160 / 198,794 | 1,304 / 1,503,761 | 0.98 (0.82-1.17) |
| Aged 80 years or older | 55 / 48,215 | 741 / 537,844 | 1.03 (0.77-1.38) |
| **Lower respiratory tract infections** | | | |
| Female | 1,972 / 334,389 | 16,738 / 1,902,866 | 0.91 (0.87-0.96) |
| Male | 2,389 / 304,387 | 19,787 / 1,759,086 | 0.89 (0.85-0.93) |
| Aged 50 to 64 years | 929 / 402,154 | 5,142 / 1,694,843 | 0.79 (0.73-0.85) |
| Aged 65 to 79 years | 1,825 / 193,449 | 14,799 / 1,466,360 | 0.92 (0.87-0.97) |
| Aged 80 years or older | 1,607 / 43,174 | 16,584 / 500,748 | 1.01 (0.96-1.07) |
| **Influenza infections** | | | |
| Female | 270 / 342,298 | 987 / 1,954,024 | 1.05 (0.91-1.21) |
| Male | 260 / 313,120 | 959 / 1,812,332 | 1.02 (0.89-1.18) |
| Aged 50 to 64 years | 138 / 406,968 | 279 / 1,716,694 | 0.99 (0.80-1.22) |
| Aged 65 to 79 years | 227 / 200,193 | 886 / 1,511,303 | 1.03 (0.89-1.20) |
| Aged 80 years or older | 165 / 48,257 | 781 / 538,359 | 1.22 (1.02-1.45) |
| **Gastrointestinal infections** | | | |
| Female | 15 / 343,352 | 76 / 1,959,560 | 1.47 (0.77-2.80) |
| Male | 9 / 314,057 | 57 / 1,817,493 | 1.03 (0.48-2.23) |
| Aged 50 to 64 years | 13 / 407,819 | 32 / 1,719,837 | 2.11 (0.93-4.79) |
| Aged 65 to 79 years | 9 / 200,918 | 63 / 1,515,810 | 1.07 (0.50-2.32) |
| Aged 80 years or older | 2 / 48,673 | 38 / 541,406 | 0.59 (0.14-2.60) |
| **Skin infections** | | | |
| Female | 756 / 337,241 | 5,351 / 1,926,404 | 0.99 (0.91-1.08) |
| Male | 1,046 / 306,220 | 6,829 / 1,774,742 | 0.97 (0.90-1.04) |
| Aged 50 to 64 years | 740 / 400,128 | 3,581 / 1,688,051 | 0.86 (0.78-0.94) |
| Aged 65 to 79 years | 683 / 196,426 | 4,947 / 1,486,196 | 1.05 (0.96-1.14) |

**Supplementary Table S1. Risk of infectious disease hospitalization after 29+ days since first SARS-CoV-2 infection in Danish 50+ year-olds and uninfected reference period during 1 January 2021 to 10 December 2022 by sex and age subgroups.**

| Outcome event / subgroup | Number of events / person-years | | Incidence rate ratio (95% CI) |
|---|---|---|---|
| | SARS-CoV-2 infected | Uninfected | |
| Aged 80 years or older | 379 / 46,907 | 3,652 / 526,900 | 1.16 (1.03-1.30) |
| **Urinary tract infections** | | | |
| Female | 718 / 337,880 | 7,080 / 1,929,220 | 0.91 (0.84-0.99) |
| Male | 790 / 310,881 | 5,897 / 1,798,928 | 1.13 (1.04-1.23) |
| Aged 50 to 64 years | 299 / 404,650 | 1,474 / 1,707,833 | 0.91 (0.79-1.06) |
| Aged 65 to 79 years | 558 / 197,910 | 4,827 / 1,496,594 | 0.96 (0.87-1.06) |
| Aged 80 years or older | 651 / 46,200 | 6,676 / 523,720 | 1.17 (1.07-1.27) |
| **Certain invasive bacterial infections** | | | |
| Female | 753 / 340,844 | 6,768 / 1,943,114 | 0.91 (0.83-0.98) |
| Male | 1,250 / 309,647 | 10,358 / 1,791,708 | 0.99 (0.93-1.05) |
| Aged 50 to 64 years | 389 / 406,046 | 2,484 / 1,711,981 | 0.72 (0.64-0.82) |
| Aged 65 to 79 years | 881 / 197,840 | 7,627 / 1,495,922 | 1.00 (0.92-1.08) |
| Aged 80 years or older | 733 / 46,605 | 7,015 / 526,919 | 1.13 (1.04-1.23) |
| **Other types of infections** | | | |
| Female | 1,075 / 336,543 | 9,270 / 1,921,986 | 0.89 (0.84-0.96) |
| Male | 1,459 / 306,347 | 10,522 / 1,775,529 | 1.01 (0.95-1.08) |
| Aged 50 to 64 years | 631 / 400,436 | 3,132 / 1,688,521 | 0.87 (0.79-0.96) |
| Aged 65 to 79 years | 1,056 / 196,318 | 8,084 / 1,485,488 | 1.02 (0.95-1.10) |
| Aged 80 years or older | 847 / 46,135 | 8,576 / 523,506 | 1.03 (0.96-1.11) |

CI denotes confidence interval.

**Supplementary Table S2. Risk of infectious disease hospitalization after 29+ days since first SARS-CoV-2 infection in Danish 50+ year-olds and uninfected reference period during 1 January 2021 to 10 December 2022 according to Covid-19 vaccination status at time of SARS-CoV-2 infection by sex and age subgroups.[a]**

| Outcome event/ vaccination status/ subgroup | Number of events / person-years | | Incidence rate ratio (95% CI) |
|---|---|---|---|
| | SARS-CoV-2 infected | Uninfected | |
| **Any infections - Unvaccinated** | | | |
| Female | 365 / 29,392 | 32,830 / 1,795,033 | 0.97 (0.87-1.08) |
| Male | 401 / 27,824 | 37,289 / 1,647,836 | 0.93 (0.84-1.03) |
| Aged 50 to 64 years | 299 / 43,628 | 12,690 / 1,606,402 | 0.84 (0.74-0.95) |
| Aged 65 to 79 years | 269 / 11,316 | 29,039 / 1,380,960 | 1.05 (0.93-1.18) |
| Aged 80 years or older | 198 / 2,273 | 28,390 / 455,507 | 1.12 (0.97-1.29) |
| **Any infections - Primary course vaccinated** | | | |
| Female | 444 / 38,030 | 26,635 / 1,306,088 | 0.95 (0.86-1.04) |
| Male | 515 / 38,318 | 29,472 / 1,166,383 | 0.91 (0.83-1.00) |
| Aged 50 to 64 years | 366 / 55,699 | 8,879 / 1,051,095 | 0.86 (0.77-0.96) |
| Aged 65 to 79 years | 370 / 17,713 | 23,146 / 1,043,067 | 1.01 (0.91-1.12) |
| Aged 80 years or older | 223 / 2,935 | 24,082 / 378,310 | 1.07 (0.94-1.23) |
| **Any infections - Booster vaccinated** | | | |
| Female | 2,877 / 242,431 | 15,183 / 731,078 | 0.85 (0.81-0.88) |
| Male | 3,664 / 214,708 | 16,616 / 654,819 | 0.91 (0.88-0.95) |
| Aged 50 to 64 years | 1,851 / 275,519 | 4,649 / 558,882 | 0.85 (0.81-0.90) |
| Aged 65 to 79 years | 2,754 / 149,784 | 12,940 / 601,144 | 0.89 (0.86-0.93) |
| Aged 80 years or older | 1,936 / 31,836 | 14,210 / 225,872 | 0.93 (0.88-0.98) |
| **Upper respiratory tract infections - Unvaccinated** | | | |
| Female | 14 / 32,181 | 1,532 / 1,941,164 | 0.59 (0.34-1.01) |
| Male | 28 / 30,775 | 1,677 / 1,801,499 | 1.28 (0.87-1.88) |
| Aged 50 to 64 years | 23 / 46,749 | 1,164 / 1,701,058 | 0.76 (0.49-1.16) |
| Aged 65 to 79 years | 13 / 12,996 | 1,304 / 1,503,761 | 1.17 (0.67-2.04) |
| Aged 80 years or older | 6 / 3,211 | 741 / 537,844 | 1.50 (0.66-3.40) |
| **Upper respiratory tract infections - Primary course vaccinated** | | | |
| Female | 47 / 41,188 | 1,202 / 1,420,986 | 1.56 (1.14-2.13) |
| Male | 36 / 41,668 | 1,232 / 1,285,977 | 1.28 (0.91-1.81) |
| Aged 50 to 64 years | 50 / 59,231 | 818 / 1,116,444 | 1.38 (1.01-1.89) |
| Aged 65 to 79 years | 23 / 19,722 | 1,012 / 1,141,360 | 1.43 (0.93-2.17) |
| Aged 80 years or older | 10 / 3,903 | 604 / 449,159 | 1.90 (1.01-3.59) |
| **Upper respiratory tract infections - Booster vaccinated** | | | |
| Female | 180 / 260,721 | 677 / 797,114 | 0.95 (0.80-1.14) |
| Male | 192 / 234,352 | 676 / 723,856 | 1.08 (0.91-1.28) |

**Supplementary Table S2. Risk of infectious disease hospitalization after 29+ days since first SARS-CoV-2 infection in Danish 50+ year-olds and uninfected reference period during 1 January 2021 to 10 December 2022 according to Covid-19 vaccination status at time of SARS-CoV-2 infection by sex and age subgroups.[a]**

| Outcome event/ vaccination status/ subgroup | Number of events / person-years | | Incidence rate ratio (95% CI) |
|---|---|---|---|
| | SARS-CoV-2 infected | Uninfected | |
| Aged 50 to 64 years | 211 / 291,030 | 425 / 594,153 | 1.13 (0.94-1.35) |
| Aged 65 to 79 years | 124 / 164,029 | 569 / 658,655 | 0.94 (0.76-1.15) |
| Aged 80 years or older | 37 / 40,015 | 359 / 268,162 | 0.85 (0.60-1.21) |
| **Lower respiratory tract infections - Unvaccinated** | | | |
| Female | 184 / 31,784 | 16,738 / 1,902,866 | 1.02 (0.88-1.18) |
| Male | 198 / 30,095 | 19,787 / 1,759,086 | 0.91 (0.79-1.05) |
| Aged 50 to 64 years | 108 / 46,704 | 5,142 / 1,694,843 | 0.77 (0.63-0.94) |
| Aged 65 to 79 years | 148 / 12,418 | 14,799 / 1,466,360 | 1.09 (0.93-1.29) |
| Aged 80 years or older | 126 / 2,757 | 16,584 / 500,748 | 1.10 (0.92-1.32) |
| **Lower respiratory tract infections - Primary course vaccinated** | | | |
| Female | 236 / 40,777 | 14,027 / 1,388,396 | 1.03 (0.91-1.18) |
| Male | 259 / 41,041 | 16,098 / 1,250,736 | 0.95 (0.84-1.08) |
| Aged 50 to 64 years | 139 / 59,317 | 3,758 / 1,111,337 | 0.83 (0.69-0.99) |
| Aged 65 to 79 years | 197 / 19,097 | 12,126 / 1,110,707 | 1.03 (0.89-1.18) |
| Aged 80 years or older | 159 / 3,404 | 14,241 / 417,088 | 1.24 (1.06-1.45) |
| **Lower respiratory tract infections - Booster vaccinated** | | | |
| Female | 1,508 / 257,139 | 8,166 / 777,865 | 0.88 (0.83-0.93) |
| Male | 1,879 / 229,784 | 9,216 / 703,272 | 0.88 (0.84-0.93) |
| Aged 50 to 64 years | 663 / 290,800 | 1,932 / 591,144 | 0.77 (0.70-0.84) |
| Aged 65 to 79 years | 1,449 / 160,003 | 6,849 / 640,767 | 0.89 (0.84-0.94) |
| Aged 80 years or older | 1,275 / 36,121 | 8,601 / 249,226 | 0.99 (0.93-1.05) |
| **Influenza infections - Unvaccinated** | | | |
| Female | 29 / 32,630 | 987 / 1,954,024 | 1.67 (1.11-2.51) |
| Male | 23 / 31,071 | 959 / 1,812,332 | 1.28 (0.82-1.99) |
| Aged 50 to 64 years | 16 / 47,404 | 279 / 1,716,694 | 1.24 (0.71-2.17) |
| Aged 65 to 79 years | 21 / 13,094 | 886 / 1,511,303 | 1.61 (1.02-2.55) |
| Aged 80 years or older | 15 / 3,202 | 781 / 538,359 | 1.94 (1.13-3.32) |
| **Influenza infections - Primary course vaccinated** | | | |
| Female | 37 / 41,731 | 955 / 1,430,094 | 1.01 (0.70-1.43) |
| Male | 35 / 42,084 | 919 / 1,293,631 | 1.13 (0.79-1.61) |
| Aged 50 to 64 years | 25 / 60,025 | 264 / 1,126,940 | 1.12 (0.71-1.77) |
| Aged 65 to 79 years | 27 / 19,878 | 851 / 1,147,193 | 1.01 (0.68-1.51) |
| Aged 80 years or older | 20 / 3,913 | 759 / 449,592 | 1.45 (0.91-2.30) |

**Supplementary Table S2. Risk of infectious disease hospitalization after 29+ days since first SARS-CoV-2 infection in Danish 50+ year-olds and uninfected reference period during 1 January 2021 to 10 December 2022 according to Covid-19 vaccination status at time of SARS-CoV-2 infection by sex and age subgroups.[a]**

| Outcome event/ vaccination status/ subgroup | Number of events / person-years | | Incidence rate ratio (95% CI) |
|---|---|---|---|
| | SARS-CoV-2 infected | Uninfected | |
| **Influenza infections - Booster vaccinated** | | | |
| Female | 199 / 263,026 | 907 / 802,059 | 0.98 (0.84-1.15) |
| Male | 200 / 236,276 | 878 / 728,065 | 0.98 (0.84-1.15) |
| Aged 50 to 64 years | 97 / 294,093 | 241 / 599,698 | 0.93 (0.73-1.19) |
| Aged 65 to 79 years | 178 / 165,151 | 813 / 661,976 | 1.00 (0.85-1.17) |
| Aged 80 years or older | 124 / 40,057 | 731 / 268,451 | 1.11 (0.91-1.35) |
| **Gastrointestinal infections - Unvaccinated** | | | |
| Female | 3 / 32,764 | 76 / 1,959,560 | 2.73 (0.76-9.80) |
| Male | 1 / 31,183 | 57 / 1,817,493 | 1.59 (0.21-12.24) |
| Aged 50 to 64 years | 4 / 47,535 | 32 / 1,719,837 | 4.06 (1.17-14.06) |
| Aged 65 to 79 years | 0 / 13,162 | 63 / 1,515,810 | NE |
| Aged 80 years or older | 0 / 3,249 | 38 / 541,406 | NE |
| **Gastrointestinal infections - Primary course vaccinated** | | | |
| Female | 3 / 41,870 | 49 / 1,434,428 | 4.14 (1.17-14.57) |
| Male | 0 / 42,208 | 40 / 1,297,679 | NE |
| Aged 50 to 64 years | 1 / 60,177 | 14 / 1,129,095 | 1.73 (0.19-15.35) |
| Aged 65 to 79 years | 2 / 19,953 | 45 / 1,150,812 | 3.10 (0.74-13.05) |
| Aged 80 years or older | 0 / 3,949 | 30 / 452,200 | NE |
| **Gastrointestinal infections - Booster vaccinated** | | | |
| Female | 9 / 263,784 | 27 / 804,590 | 1.10 (0.50-2.45) |
| Male | 8 / 236,965 | 30 / 730,456 | 1.10 (0.48-2.51) |
| Aged 50 to 64 years | 8 / 294,647 | 9 / 600,838 | 1.81 (0.66-4.96) |
| Aged 65 to 79 years | 7 / 165,721 | 32 / 664,165 | 0.96 (0.41-2.27) |
| Aged 80 years or older | 2 / 40,381 | 16 / 270,043 | 0.71 (0.16-3.21) |
| **Skin infections - Unvaccinated** | | | |
| Female | 75 / 32,075 | 5,351 / 1,926,404 | 1.03 (0.81-1.30) |
| Male | 111 / 30,307 | 6,829 / 1,774,742 | 1.07 (0.88-1.31) |
| Aged 50 to 64 years | 92 / 46,562 | 3,581 / 1,688,051 | 0.86 (0.69-1.07) |
| Aged 65 to 79 years | 60 / 12,756 | 4,947 / 1,486,196 | 1.27 (0.97-1.64) |
| Aged 80 years or older | 34 / 3,065 | 3,652 / 526,900 | 1.38 (0.98-1.95) |
| **Skin infections - Primary course vaccinated** | | | |
| Female | 81 / 41,065 | 4,183 / 1,408,975 | 0.95 (0.76-1.20) |
| Male | 124 / 41,148 | 5,244 / 1,265,468 | 0.93 (0.77-1.12) |

**Supplementary Table S2. Risk of infectious disease hospitalization after 29+ days since first SARS-CoV-2 infection in Danish 50+ year-olds and uninfected reference period during 1 January 2021 to 10 December 2022 according to Covid-19 vaccination status at time of SARS-CoV-2 infection by sex and age subgroups.[a]**

| Outcome event/ vaccination status/ subgroup | Number of events / person-years | | Incidence rate ratio (95% CI) |
|---|---|---|---|
| | SARS-CoV-2 infected | Uninfected | |
| Aged 50 to 64 years | 98 / 58,972 | 2,400 / 1,107,258 | 0.79 (0.64-0.98) |
| Aged 65 to 79 years | 73 / 19,464 | 3,948 / 1,127,392 | 1.09 (0.86-1.38) |
| Aged 80 years or older | 34 / 3,777 | 3,079 / 439,793 | 1.25 (0.88-1.76) |
| **Skin infections - Booster vaccinated** | | | |
| Female | 581 / 259,293 | 2,286 / 790,045 | 0.99 (0.89-1.09) |
| Male | 800 / 231,192 | 2,875 / 711,948 | 0.97 (0.89-1.05) |
| Aged 50 to 64 years | 538 / 289,270 | 1,262 / 588,974 | 0.88 (0.79-0.98) |
| Aged 65 to 79 years | 540 / 162,191 | 2,164 / 650,444 | 1.01 (0.92-1.12) |
| Aged 80 years or older | 303 / 39,024 | 1,735 / 262,574 | 1.13 (0.99-1.28) |
| **Urinary tract infections - Unvaccinated** | | | |
| Female | 88 / 32,116 | 7,080 / 1,929,220 | 1.15 (0.92-1.43) |
| Male | 68 / 30,890 | 5,897 / 1,798,928 | 1.15 (0.90-1.47) |
| Aged 50 to 64 years | 33 / 47,106 | 1,474 / 1,707,833 | 0.85 (0.59-1.22) |
| Aged 65 to 79 years | 43 / 12,886 | 4,827 / 1,496,594 | 1.01 (0.75-1.38) |
| Aged 80 years or older | 80 / 3,014 | 6,676 / 523,720 | 1.58 (1.26-1.99) |
| **Urinary tract infections - Primary course vaccinated** | | | |
| Female | 101 / 41,134 | 5,673 / 1,410,385 | 1.14 (0.93-1.39) |
| Male | 86 / 41,831 | 4,675 / 1,282,551 | 1.13 (0.91-1.41) |
| Aged 50 to 64 years | 47 / 59,662 | 1,000 / 1,120,786 | 0.99 (0.72-1.35) |
| Aged 65 to 79 years | 67 / 19,586 | 3,778 / 1,135,300 | 1.17 (0.92-1.50) |
| Aged 80 years or older | 73 / 3,716 | 5,570 / 436,851 | 1.34 (1.06-1.70) |
| **Urinary tract infections - Booster vaccinated** | | | |
| Female | 499 / 259,874 | 2,954 / 790,893 | 0.81 (0.74-0.90) |
| Male | 610 / 234,546 | 2,484 / 721,691 | 1.10 (1.00-1.21) |
| Aged 50 to 64 years | 210 / 292,506 | 513 / 596,401 | 0.89 (0.75-1.05) |
| Aged 65 to 79 years | 433 / 163,421 | 2,028 / 655,217 | 0.93 (0.83-1.03) |
| Aged 80 years or older | 466 / 38,493 | 2,897 / 260,965 | 1.06 (0.96-1.17) |
| **Certain invasive bacterial infections - Unvaccinated** | | | |
| Female | 71 / 32,498 | 6,768 / 1,943,114 | 0.95 (0.75-1.21) |
| Male | 144 / 30,717 | 10,358 / 1,791,708 | 1.40 (1.18-1.66) |
| Aged 50 to 64 years | 53 / 47,310 | 2,484 / 1,711,981 | 0.86 (0.65-1.15) |
| Aged 65 to 79 years | 90 / 12,860 | 7,627 / 1,495,922 | 1.36 (1.10-1.69) |
| Aged 80 years or older | 72 / 3,045 | 7,015 / 526,919 | 1.54 (1.21-1.95) |

**Supplementary Table S2. Risk of infectious disease hospitalization after 29+ days since first SARS-CoV-2 infection in Danish 50+ year-olds and uninfected reference period during 1 January 2021 to 10 December 2022 according to Covid-19 vaccination status at time of SARS-CoV-2 infection by sex and age subgroups.[a]**

| Outcome event/ vaccination status/ subgroup | Number of events / person-years | | Incidence rate ratio (95% CI) |
|---|---|---|---|
| | SARS-CoV-2 infected | Uninfected | |
| **Certain invasive bacterial infections - Primary course vaccinated** | | | |
| Female | 102 / 41,544 | 5,414 / 1,420,848 | 1.13 (0.92-1.38) |
| Male | 143 / 41,723 | 8,199 / 1,276,542 | 1.03 (0.87-1.22) |
| Aged 50 to 64 years | 74 / 59,895 | 1,790 / 1,123,307 | 0.94 (0.73-1.20) |
| Aged 65 to 79 years | 100 / 19,628 | 5,951 / 1,134,460 | 1.12 (0.91-1.37) |
| Aged 80 years or older | 71 / 3,744 | 5,872 / 439,623 | 1.32 (1.04-1.67) |
| **Certain invasive bacterial infections - Booster vaccinated** | | | |
| Female | 556 / 261,955 | 2,918 / 796,648 | 0.85 (0.77-0.94) |
| Male | 930 / 233,604 | 4,318 / 718,138 | 0.93 (0.87-1.00) |
| Aged 50 to 64 years | 255 / 293,417 | 898 / 597,627 | 0.65 (0.56-0.75) |
| Aged 65 to 79 years | 668 / 163,339 | 3,120 / 654,580 | 0.94 (0.86-1.02) |
| Aged 80 years or older | 563 / 38,803 | 3,218 / 262,579 | 1.07 (0.98-1.18) |
| **Other types of infections - Unvaccinated** | | | |
| Female | 118 / 31,938 | 9,270 / 1,921,986 | 1.03 (0.85-1.24) |
| Male | 142 / 30,239 | 10,522 / 1,775,529 | 1.11 (0.94-1.32) |
| Aged 50 to 64 years | 89 / 46,425 | 3,132 / 1,688,521 | 1.02 (0.82-1.28) |
| Aged 65 to 79 years | 84 / 12,741 | 8,084 / 1,485,488 | 1.07 (0.86-1.34) |
| Aged 80 years or older | 87 / 3,010 | 8,576 / 523,506 | 1.27 (1.03-1.58) |
| **Other types of infections - Primary course vaccinated** | | | |
| Female | 138 / 40,939 | 7,507 / 1,405,386 | 1.07 (0.90-1.27) |
| Male | 158 / 41,123 | 8,417 / 1,265,619 | 1.00 (0.85-1.18) |
| Aged 50 to 64 years | 89 / 58,956 | 2,206 / 1,107,681 | 0.91 (0.73-1.14) |
| Aged 65 to 79 years | 116 / 19,407 | 6,419 / 1,126,560 | 1.16 (0.96-1.40) |
| Aged 80 years or older | 91 / 3,698 | 7,299 / 436,764 | 1.22 (0.99-1.51) |
| **Other types of infections - Booster vaccinated** | | | |
| Female | 789 / 258,896 | 4,250 / 787,974 | 0.85 (0.78-0.92) |
| Male | 1,127 / 231,476 | 4,682 / 712,067 | 0.99 (0.93-1.07) |
| Aged 50 to 64 years | 437 / 289,755 | 1,159 / 589,416 | 0.82 (0.73-0.92) |
| Aged 65 to 79 years | 836 / 162,183 | 3,574 / 649,925 | 1.00 (0.93-1.08) |
| Aged 80 years or older | 643 / 38,434 | 4,199 / 260,700 | 0.97 (0.89-1.06) |

[a]Unvaccinated (zero doses), primary course vaccinated (2 doses), and booster (3 or 4 doses) denotes the number Covid-19 vaccines received at time of the first SARS-CoV-2 infection. CI denotes confidence interval and NE not estimable.

**Supplementary Table S3. Sensitivity analysis of risk of infectious disease hospitalization where using an alternative main risk period of ≥90+ days since first SARS-CoV-2 infection.**

| Outcome event/subgroup | Number of events / person-years | | Incidence rate ratio (95% CI) |
|---|---|---|---|
| | SARS-CoV-2 infected | Uninfected | |
| **Any infections** | | | |
| All | 6,150 / 459,558 | 70,119 / 3,442,869 | 0.86 (0.84-0.89) |
| Female | 2,803 / 241,009 | 32,830 / 1,795,033 | 0.86 (0.83-0.90) |
| Male | 3,347 / 218,549 | 37,289 / 1,647,836 | 0.86 (0.83-0.90) |
| Aged 50 to 64 years | 1,948 / 294,218 | 12,690 / 1,606,402 | 0.83 (0.79-0.88) |
| Aged 65 to 79 years | 2,481 / 136,997 | 29,039 / 1,380,960 | 0.88 (0.84-0.92) |
| Aged 80 years or older | 1,721 / 28,343 | 28,390 / 455,507 | 0.91 (0.87-0.96) |
| **Upper respiratory tract infections** | | | |
| All | 373 / 498,980 | 3,209 / 3,742,663 | 1.07 (0.95-1.21) |
| Female | 180 / 260,088 | 1,532 / 1,941,164 | 0.98 (0.82-1.17) |
| Male | 193 / 238,892 | 1,677 / 1,801,499 | 1.16 (0.98-1.37) |
| Aged 50 to 64 years | 217 / 311,863 | 1,164 / 1,701,058 | 1.07 (0.90-1.28) |
| Aged 65 to 79 years | 114 / 150,928 | 1,304 / 1,503,761 | 0.96 (0.77-1.18) |
| Aged 80 years or older | 42 / 36,189 | 741 / 537,844 | 1.18 (0.84-1.65) |
| **Lower respiratory tract infections** | | | |
| All | 3,140 / 490,998 | 36,525 / 3,661,952 | 0.84 (0.81-0.87) |
| Female | 1,440 / 256,660 | 16,738 / 1,902,866 | 0.86 (0.81-0.91) |
| Male | 1,700 / 234,338 | 19,787 / 1,759,086 | 0.82 (0.78-0.87) |
| Aged 50 to 64 years | 711 / 311,705 | 5,142 / 1,694,843 | 0.76 (0.70-0.83) |
| Aged 65 to 79 years | 1,301 / 146,869 | 14,799 / 1,466,360 | 0.85 (0.80-0.91) |
| Aged 80 years or older | 1,128 / 32,424 | 16,584 / 500,748 | 0.93 (0.87-0.99) |
| **Influenza infections** | | | |
| All | 187 / 503,660 | 1,946 / 3,766,356 | 1.22 (1.04-1.42) |
| Female | 97 / 262,684 | 987 / 1,954,024 | 1.25 (1.01-1.56) |
| Male | 90 / 240,976 | 959 / 1,812,332 | 1.18 (0.94-1.47) |
| Aged 50 to 64 years | 46 / 315,451 | 279 / 1,716,694 | 0.93 (0.66-1.29) |
| Aged 65 to 79 years | 84 / 151,990 | 886 / 1,511,303 | 1.32 (1.05-1.67) |
| Aged 80 years or older | 57 / 36,219 | 781 / 538,359 | 1.54 (1.17-2.03) |
| **Gastrointestinal infections** | | | |
| All | 19 / 505,199 | 133 / 3,777,053 | 1.28 (0.74-2.21) |
| Female | 11 / 263,504 | 76 / 1,959,560 | 1.32 (0.64-2.75) |
| Male | 8 / 241,696 | 57 / 1,817,493 | 1.20 (0.52-2.75) |
| Aged 50 to 64 years | 12 / 316,117 | 32 / 1,719,837 | 2.49 (1.03-6.03) |
| Aged 65 to 79 years | 7 / 152,548 | 63 / 1,515,810 | 1.09 (0.46-2.58) |
| Aged 80 years or older | 0 / 36,534 | 38 / 541,406 | 0.00 (0.00-Inf) |
| **Skin infections** | | | |

**Supplementary Table S3. Sensitivity analysis of risk of infectious disease hospitalization where using an alternative main risk period of ≥90+ days since first SARS-CoV-2 infection.**

| Outcome event/subgroup | Number of events / person-years | | Incidence rate ratio (95% CI) |
|---|---|---|---|
| | SARS-CoV-2 infected | Uninfected | |
| All | 1,373 / 494,409 | 12,180 / 3,701,146 | 0.96 (0.91-1.03) |
| Female | 580 / 258,781 | 5,351 / 1,926,404 | 0.99 (0.90-1.09) |
| Male | 793 / 235,628 | 6,829 / 1,774,742 | 0.95 (0.87-1.03) |
| Aged 50 to 64 years | 572 / 310,111 | 3,581 / 1,688,051 | 0.84 (0.76-0.93) |
| Aged 65 to 79 years | 522 / 149,105 | 4,947 / 1,486,196 | 1.04 (0.94-1.15) |
| Aged 80 years or older | 279 / 35,193 | 3,652 / 526,900 | 1.14 (1.00-1.30) |
| **Urinary tract infections** | | | |
| All | 1,104 / 498,559 | 12,977 / 3,728,148 | 0.98 (0.91-1.05) |
| Female | 536 / 259,294 | 7,080 / 1,929,220 | 0.89 (0.81-0.98) |
| Male | 568 / 239,265 | 5,897 / 1,798,928 | 1.07 (0.98-1.18) |
| Aged 50 to 64 years | 232 / 313,650 | 1,474 / 1,707,833 | 0.89 (0.76-1.05) |
| Aged 65 to 79 years | 411 / 150,243 | 4,827 / 1,496,594 | 0.92 (0.83-1.03) |
| Aged 80 years or older | 461 / 34,666 | 6,676 / 523,720 | 1.12 (1.01-1.24) |
| **Certain invasive bacterial infections** | | | |
| All | 1,409 / 499,920 | 17,126 / 3,734,822 | 0.87 (0.82-0.93) |
| Female | 564 / 261,583 | 6,768 / 1,943,114 | 0.89 (0.81-0.98) |
| Male | 845 / 238,337 | 10,358 / 1,791,708 | 0.86 (0.80-0.93) |
| Aged 50 to 64 years | 281 / 314,743 | 2,484 / 1,711,981 | 0.67 (0.58-0.76) |
| Aged 65 to 79 years | 604 / 150,201 | 7,627 / 1,495,922 | 0.89 (0.81-0.97) |
| Aged 80 years or older | 524 / 34,976 | 7,015 / 526,919 | 1.07 (0.97-1.18) |
| **Other types of infections** | | | |
| All | 1,853 / 494,030 | 19,792 / 3,697,515 | 0.90 (0.86-0.95) |
| Female | 798 / 258,275 | 9,270 / 1,921,986 | 0.85 (0.79-0.92) |
| Male | 1,055 / 235,756 | 10,522 / 1,775,529 | 0.95 (0.88-1.02) |
| Aged 50 to 64 years | 493 / 310,360 | 3,132 / 1,688,521 | 0.85 (0.76-0.94) |
| Aged 65 to 79 years | 749 / 149,041 | 8,084 / 1,485,488 | 0.94 (0.86-1.02) |
| Aged 80 years or older | 611 / 34,630 | 8,576 / 523,506 | 0.98 (0.90-1.07) |

CI denotes confidence interval and NE not estimable.

**Supplementary Table S4. Sensitivity analysis of risk of infectious disease hospitalization excluding a 7-day pre-risk period prior to the first SARS-CoV-2 infection.**

| Outcome event / subgroup | Number of events / person-years | | Incidence rate ratio (95% CI) |
|---|---|---|---|
| | SARS-CoV-2 infected | Uninfected[a] | |
| **Any infections** | | | |
| All | 8,436 / 598,038 | 68,045 / 3,426,907 | 0.93 (0.91-0.95) |
| Female | 3,771 / 314,099 | 31,956 / 1,786,620 | 0.91 (0.88-0.94) |
| Male | 4,665 / 283,940 | 36,089 / 1,640,287 | 0.95 (0.92-0.98) |
| Aged 50 to 64 years | 2,568 / 379,743 | 12,456 / 1,596,634 | 0.87 (0.83-0.92) |
| Aged 65 to 79 years | 3,444 / 180,538 | 28,237 / 1,375,918 | 0.95 (0.91-0.99) |
| Aged 80 years or older | 2,424 / 37,758 | 27,352 / 454,354 | 1.00 (0.96-1.05) |
| **Upper respiratory tract infections** | | | |
| All | 507 / 649,374 | 3,083 / 3,725,327 | 1.13 (1.01-1.25) |
| Female | 245 / 338,938 | 1,470 / 1,932,086 | 1.04 (0.89-1.21) |
| Male | 262 / 310,435 | 1,613 / 1,793,241 | 1.21 (1.04-1.40) |
| Aged 50 to 64 years | 292 / 402,365 | 1,139 / 1,690,727 | 1.15 (0.99-1.34) |
| Aged 65 to 79 years | 160 / 198,794 | 1,250 / 1,498,226 | 1.03 (0.86-1.23) |
| Aged 80 years or older | 55 / 48,215 | 694 / 536,375 | 1.12 (0.83-1.50) |
| **Lower respiratory tract infections** | | | |
| All | 4,361 / 638,777 | 35,225 / 3,644,922 | 0.94 (0.91-0.97) |
| Female | 1,972 / 334,389 | 16,224 / 1,893,920 | 0.94 (0.90-0.99) |
| Male | 2,389 / 304,387 | 19,001 / 1,751,002 | 0.93 (0.89-0.98) |
| Aged 50 to 64 years | 929 / 402,154 | 5,027 / 1,684,518 | 0.80 (0.74-0.87) |
| Aged 65 to 79 years | 1,825 / 193,449 | 14,296 / 1,460,970 | 0.96 (0.91-1.01) |
| Aged 80 years or older | 1,607 / 43,174 | 15,902 / 499,433 | 1.06 (1.01-1.12) |
| **Influenza infections** | | | |
| All | 530 / 655,418 | 1,877 / 3,748,864 | 1.06 (0.96-1.18) |
| Female | 270 / 342,298 | 950 / 1,944,859 | 1.08 (0.94-1.24) |
| Male | 260 / 313,120 | 927 / 1,804,005 | 1.04 (0.91-1.20) |
| Aged 50 to 64 years | 138 / 406,968 | 271 / 1,706,248 | 1.01 (0.82-1.24) |
| Aged 65 to 79 years | 227 / 200,193 | 855 / 1,505,729 | 1.06 (0.91-1.22) |
| Aged 80 years or older | 165 / 48,257 | 751 / 536,888 | 1.26 (1.06-1.49) |
| **Gastrointestinal infections** | | | |
| All | 24 / 657,409 | 130 / 3,759,512 | 1.31 (0.80-2.15) |
| Female | 15 / 343,352 | 74 / 1,950,369 | 1.54 (0.81-2.94) |
| Male | 9 / 314,057 | 56 / 1,809,143 | 1.04 (0.48-2.27) |
| Aged 50 to 64 years | 13 / 407,819 | 31 / 1,709,371 | 2.21 (0.97-5.07) |
| Aged 65 to 79 years | 9 / 200,918 | 63 / 1,510,218 | 1.07 (0.50-2.31) |
| Aged 80 years or older | 2 / 48,673 | 36 / 539,923 | 0.63 (0.14-2.78) |
| **Skin infections** | | | |

**Supplementary Table S4. Sensitivity analysis of risk of infectious disease hospitalization excluding a 7-day pre-risk period prior to the first SARS-CoV-2 infection.**

| Outcome event / subgroup | Number of events / person-years | | Incidence rate ratio (95% CI) |
|---|---|---|---|
| | SARS-CoV-2 infected | Uninfected[a] | |
| All | 1,802 / 643,461 | 11,997 / 3,683,969 | 0.99 (0.94-1.05) |
| Female | 756 / 337,241 | 5,280 / 1,917,374 | 1.01 (0.93-1.10) |
| Male | 1,046 / 306,220 | 6,717 / 1,766,595 | 0.99 (0.92-1.06) |
| Aged 50 to 64 years | 740 / 400,128 | 3,541 / 1,677,776 | 0.86 (0.79-0.95) |
| Aged 65 to 79 years | 683 / 196,426 | 4,875 / 1,480,724 | 1.06 (0.97-1.16) |
| Aged 80 years or older | 379 / 46,907 | 3,581 / 525,469 | 1.18 (1.06-1.33) |
| **Urinary tract infections** | | | |
| All | 1,508 / 648,761 | 12,611 / 3,710,841 | 1.05 (0.99-1.11) |
| Female | 718 / 337,880 | 6,880 / 1,920,176 | 0.94 (0.87-1.03) |
| Male | 790 / 310,881 | 5,731 / 1,790,664 | 1.17 (1.08-1.27) |
| Aged 50 to 64 years | 299 / 404,650 | 1,443 / 1,697,447 | 0.93 (0.81-1.08) |
| Aged 65 to 79 years | 558 / 197,910 | 4,710 / 1,491,082 | 0.99 (0.90-1.09) |
| Aged 80 years or older | 651 / 46,200 | 6,458 / 522,312 | 1.22 (1.12-1.33) |
| **Certain invasive bacterial infections** | | | |
| All | 2,003 / 650,491 | 16,656 / 3,717,472 | 0.99 (0.94-1.04) |
| Female | 753 / 340,844 | 6,601 / 1,933,993 | 0.93 (0.86-1.01) |
| Male | 1,250 / 309,647 | 10,055 / 1,783,480 | 1.02 (0.96-1.09) |
| Aged 50 to 64 years | 389 / 406,046 | 2,427 / 1,701,559 | 0.74 (0.66-0.83) |
| Aged 65 to 79 years | 881 / 197,840 | 7,434 / 1,490,414 | 1.03 (0.95-1.11) |
| Aged 80 years or older | 733 / 46,605 | 6,795 / 525,499 | 1.18 (1.08-1.28) |
| **Other types of infections** | | | |
| All | 2,534 / 642,890 | 19,330 / 3,680,361 | 0.98 (0.94-1.03) |
| Female | 1,075 / 336,543 | 9,087 / 1,912,977 | 0.91 (0.85-0.98) |
| Male | 1,459 / 306,347 | 10,243 / 1,767,383 | 1.04 (0.98-1.11) |
| Aged 50 to 64 years | 631 / 400,436 | 3,081 / 1,678,240 | 0.88 (0.80-0.97) |
| Aged 65 to 79 years | 1,056 / 196,318 | 7,915 / 1,480,022 | 1.05 (0.97-1.12) |
| Aged 80 years or older | 847 / 46,135 | 8,334 / 522,100 | 1.07 (0.99-1.15) |

[a]The pre-risk period of the last -7 days before the first SARS-CoV-2 infection date were omitted from the reference period. CI denotes confidence interval.

**Supplementary Table S5. Risk of infectious disease hospitalization with tonsillitis after after 29-180 days and >180 days since first SARS-CoV-2 infection during 1 January 2021 to 10 December 2022.**

| | Number of events / person-years | | | Incidence rate ratio (95% CI) | |
|---|---|---|---|---|---|
| | SARS-CoV-2 infected | | Uninfected | 29-180 days | >180 days |
| | 29-180 days | >180 days | | | |
| All | 44 / 360,133 | 35 / 295,854 | 336 / 3,772,194 | 1.25 (0.88-1.79) | 1.45 (0.96-2.19) |
| Female | 29 / 188,913 | 17 / 153,534 | 183 / 1,956,579 | 1.30 (0.83-2.03) | 1.24 (0.69-2.22) |
| Male | 15 / 171,220 | 18 / 142,320 | 153 / 1,815,615 | 1.13 (0.63-2.03) | 1.71 (0.95-3.08) |
| Aged 50 to 64 years | 31 / 218,421 | 27 / 188,219 | 185 / 1,716,297 | 1.23 (0.79-1.91) | 1.51 (0.92-2.50) |
| Aged 65 to 79 years | 11 / 113,651 | 7 / 87,053 | 111 / 1,514,703 | 1.25 (0.63-2.45) | 1.38 (0.58-3.27) |
| Aged 80 years or older | 2 / 28,061 | 1 / 20,582 | 40 / 541,194 | 1.18 (0.26-5.34) | 0.62 (0.08-4.97) |

CI denotes confidence interval.

**Supplementary Table S6. Risk of infectious disease hospitalization after 90+ days since first Covid-19 hospitalization in Danish 50+ year-olds and uninfected reference period during 1 January 2021 to 10 December 2022.[a]**

| Outcome event / subgroup | Number of events / person-years | | Incidence rate ratio (95% CI) |
|---|---|---|---|
| | Covid-19 hospitalized | Uninfected | |
| **Any infections** | | | |
| All | 488 / 6,375 | 77,034 / 4,095,436 | 2.28 (2.08-2.49) |
| Female | 204 / 3,007 | 36,004 / 2,138,410 | 2.28 (1.99-2.62) |
| Male | 284 / 3,368 | 41,030 / 1,957,026 | 2.27 (2.02-2.55) |
| Aged 50 to 64 years | 77 / 2,577 | 15,166 / 2,021,827 | 2.56 (2.04-3.20) |
| Aged 65 to 79 years | 188 / 2,326 | 31,923 / 1,578,203 | 2.59 (2.24-2.99) |
| Aged 80 years or older | 223 / 1,472 | 29,945 / 495,406 | 1.95 (1.71-2.22) |
| **Upper respiratory tract infections** | | | |
| All | 23 / 10,220 | 3,624 / 4,446,339 | 1.97 (1.31-2.98) |
| Female | 5 / 4,634 | 1,731 / 2,309,622 | 0.99 (0.41-2.40) |
| Male | 18 / 5,587 | 1,893 / 2,136,716 | 2.73 (1.71-4.36) |
| Aged 50 to 64 years | 5 / 3,407 | 1,445 / 2,140,208 | 1.42 (0.59-3.43) |
| Aged 65 to 79 years | 10 / 3,831 | 1,429 / 1,719,005 | 2.33 (1.25-4.36) |
| Aged 80 years or older | 8 / 2,983 | 750 / 587,125 | 2.32 (1.15-4.70) |
| **Influenza infections** | | | |
| All | 33 / 10,300 | 2,367 / 4,476,595 | 2.79 (1.98-3.95) |
| Female | 14 / 4,676 | 1,203 / 2,326,131 | 2.64 (1.55-4.49) |
| Male | 19 / 5,624 | 1,164 / 2,150,465 | 2.92 (1.85-4.62) |
| Aged 50 to 64 years | 11 / 3,475 | 398 / 2,160,815 | 5.99 (3.25-11.06) |
| Aged 65 to 79 years | 11 / 3,860 | 1,065 / 1,728,064 | 2.30 (1.26-4.18) |
| Aged 80 years or older | 11 / 2,965 | 904 / 587,716 | 2.06 (1.13-3.75) |
| **Lower respiratory tract infections** | | | |
| All | 357 / 8,199 | 39,590 / 4,356,870 | 2.33 (2.10-2.59) |
| Female | 142 / 3,814 | 18,226 / 2,267,490 | 2.31 (1.95-2.72) |
| Male | 215 / 4,385 | 21,364 / 2,089,381 | 2.34 (2.04-2.68) |
| Aged 50 to 64 years | 53 / 3,102 | 5,950 / 2,134,194 | 3.26 (2.48-4.28) |
| Aged 65 to 79 years | 137 / 3,005 | 16,138 / 1,676,905 | 2.74 (2.32-3.25) |
| Aged 80 years or older | 167 / 2,092 | 17,502 / 545,771 | 1.86 (1.60-2.17) |
| **Gastrointestinal infections** | | | |
| All | 2 / 10,506 | 154 / 4,489,178 | 2.99 (0.73-12.31) |
| Female | 1 / 4,776 | 89 / 2,332,679 | 3.01 (0.41-22.16) |
| Male | 1 / 5,730 | 65 / 2,156,499 | 2.91 (0.39-21.55) |
| Aged 50 to 64 years | 1 / 3,525 | 44 / 2,164,819 | 6.83 (0.89-52.37) |
| Aged 65 to 79 years | 1 / 3,937 | 71 / 1,733,254 | 4.08 (0.55-30.18) |
| Aged 80 years or older | 0 / 3,044 | 39 / 591,105 | NE |

**Supplementary Table S6. Risk of infectious disease hospitalization after 90+ days since first Covid-19 hospitalization in Danish 50+ year-olds and uninfected reference period during 1 January 2021 to 10 December 2022.[a]**

| Outcome event / subgroup | Number of events / person-years | | Incidence rate ratio (95% CI) |
|---|---|---|---|
| | Covid-19 hospitalized | Uninfected | |
| **Skin infections** | | | |
| All | 113 / 9,841 | 13,881 / 4,398,842 | 2.04 (1.69-2.46) |
| Female | 50 / 4,516 | 6,066 / 2,293,151 | 2.39 (1.80-3.16) |
| Male | 63 / 5,325 | 7,815 / 2,105,691 | 1.85 (1.44-2.37) |
| Aged 50 to 64 years | 22 / 3,328 | 4,338 / 2,124,852 | 2.05 (1.34-3.12) |
| Aged 65 to 79 years | 44 / 3,685 | 5,585 / 1,699,036 | 2.23 (1.65-3.00) |
| Aged 80 years or older | 47 / 2,827 | 3,958 / 574,954 | 1.92 (1.44-2.57) |
| **Urinary tract infections** | | | |
| All | 157 / 9,740 | 14,143 / 4,431,809 | 2.42 (2.06-2.83) |
| Female | 67 / 4,376 | 7,621 / 2,296,864 | 2.22 (1.74-2.83) |
| Male | 90 / 5,364 | 6,522 / 2,134,945 | 2.57 (2.08-3.17) |
| Aged 50 to 64 years | 23 / 3,405 | 1,746 / 2,149,497 | 4.53 (2.98-6.89) |
| Aged 65 to 79 years | 42 / 3,664 | 5,295 / 1,711,108 | 2.17 (1.60-2.95) |
| Aged 80 years or older | 92 / 2,672 | 7,102 / 571,204 | 2.22 (1.80-2.73) |
| **Certain invasive bacterial infections** | | | |
| All | 198 / 9,662 | 18,628 / 4,440,520 | 2.40 (2.09-2.77) |
| Female | 82 / 4,483 | 7,333 / 2,313,880 | 2.86 (2.30-3.56) |
| Male | 116 / 5,178 | 11,295 / 2,126,639 | 2.15 (1.79-2.59) |
| Aged 50 to 64 years | 16 / 3,372 | 2,829 / 2,155,231 | 1.86 (1.13-3.05) |
| Aged 65 to 79 years | 81 / 3,593 | 8,314 / 1,710,463 | 2.79 (2.23-3.47) |
| Aged 80 years or older | 101 / 2,697 | 7,485 / 574,826 | 2.19 (1.79-2.67) |
| **Other types of infections** | | | |
| All | 262 / 9,439 | 21,847 / 4,395,163 | 2.50 (2.21-2.82) |
| Female | 102 / 4,356 | 10,184 / 2,288,195 | 2.38 (1.95-2.90) |
| Male | 160 / 5,082 | 11,663 / 2,106,968 | 2.58 (2.20-3.02) |
| Aged 50 to 64 years | 43 / 3,258 | 3,704 / 2,125,756 | 3.65 (2.69-4.94) |
| Aged 65 to 79 years | 96 / 3,524 | 8,959 / 1,698,446 | 2.79 (2.28-3.42) |
| Aged 80 years or older | 123 / 2,657 | 9,184 / 570,961 | 2.03 (1.70-2.43) |

[a]First Covid-19 hospitalization was defined as the first inpatient hospitalization during the study period with a registered primary or secondary diagnoses for Covid-19 (International Classification of Diseases System, version 10-codes: U071, B342, B948A, B972, Z038PA1) and a recorded positive PCR test for SARS-CoV-2 within 14 days before to 2 days after the admission date (the admission date served as the index date). The main risk period was ≥day 90 following the index date.

**Supplementary Table S7. Eligibility criteria and exposure and covariates definitions.**

| Variable | Details |
|---|---|
| **Eligibility criteria** | |
| Age of ≥50 years | *The Civil Registration System*[1]. The register provides the mandatory unique personal identifier for all permanent residents of Denmark allowing the cross-linkage of all Danish healthcare services and civil registrations systems. The register also holds demographic information such as birthdate, sex, continuously updated information and dates on historical addresses, immigration and emigration status, and death. Age was defined by year of study entry minus birthyear. |
| Danish residency | *The Civil Registration System*[1]. Defined as known Danish address (and a unique personal identifier) at baseline (start of the study period, 1 January 2021). |
| Initially uninfected with SARS-CoV-2 prior study entry | *The Danish Microbiology Database*[2]. The register holds information on all microbiology samples analyzed at Danish departments of microbiology, including information on SARS-CoV-2 PCR test results, date of sampling, date of analysis, type of test, and interpretation of test. PCR tests for SARS-CoV-2 were freely available to all individuals in Denmark regardless of testing reason during the study period. Uninfected prior to start of study period was defined as no past records of positive PCR test results for SARS-CoV-2. |
| **Exposure** | |
| SARS-CoV-2 infection | *The Danish Microbiology Database*[2]. Defined by the first positive PCR test result for SARS-CoV-2 during the study period. The testing date served as the index date (that is, day zero), and exposure status was handled as a time-varying exposure variable. The main risk period of was ≥29 days (4 weeks) since the index date and onwards, that is, we considered the period to be post the symptomatic acute phase of SARS-CoV-2. |
| **Covariates** | |
| *Time-fixed covariates (at baseline)* | |
| Sex | *The Civil Registration System*[1]. Defined by registered sex. |
| Ethnicity | *The Civil Registration System*[1]. Defined by registered place of birth and categorized according Nordic (Denmark, Finland, Norway, or Sweden), Western (rest of Europe, united states, Australia or New Zealand), and non-Western countries (all others). |
| Region of residency | *The Civil Registration System*[1]. Defined by the last registered address and categorized according: Northern Denmark Region, Central Denmark Region, Region of Southern Denmark, Capital Region of Denmark, and Region Zealand. |

| | |
|---|---|
| Vaccination priority groups | *The Danish Vaccination Register*[3]. The register holds information on all (mandatorily) recorded administered vaccines in Denmark including information on vaccination date, -type, -dose, and -product batch. In addition, during the Covid-19 pandemic, the register was allocated information on governmentally prioritized Covid-19 vaccine groups assigned according to whether an individual was considered as being at high risk of severe Covid-19 (categorized binarily). |
| Comorbidities | *The National Patient Register*[4]*.* The register holds information on all hospital contacts (secondary healthcare facilities) in Denmark including information on the contact duration and treating physician-assigned diagnoses (registered according to the International Classification of Diseases System, revision 10-codes). We defined comorbidity status as any registered primary or secondary diagnosis regardless of the hospital contact type between 1 January 2018 and baseline (1 January 2021) and categorized the number of comorbidities as a sum (0, 1, or ≥2 comorbidities). |
| Asthma | ICD-10 codes: J45, J46 |
| Chronic respiratory disorder | ICD-10 codes: E84, J41-J44, J47, J84 |
| Chronic cardiac disorder | ICD-10 codes: I05-I08, I20-I28, I34-I37, I42-I51 |
| Renal disorder | ICD-10 codes: N03, N05, N07, N18, N19, N25-N27 |
| Diabetes | ICD-10 codes: E10-E14 |
| Autoimmune disorder | ICD-10 codes: D510, D590, D591, D690, D693, D86, E035, E039, E050, E055, E059, E063, E065, E271, E272, E310, G04, G131, G35, G36, G61, G700, H20, I00, I02, K50, K51, K732, K743, K900, L10, L12, L130, L40, L63, L80, M05, M06, M08, M30, M311, M313, M315, M316, M317, M32-M34, M350-M353, M358, M359, M45, M60 |
| Epilepsy | ICD-10 codes: G40, G41 |
| Malignancy | ICD-10 codes: C00-C96 (not C44), D70-D72, D730, D81-D84 |
| Psychiatric disorder | ICD-10 codes: F00-F99 |
| *Time-varying covariates* | |
| Age | *The Civil Registration System*[1]. Age was defined by year of study (i.e., year 2021 or 2022) minus birthyear and categorized in 5-year bins (9 levels: 50-54, 55-59, 60-64, 65-69, 70-74, 75-79, 80-84, 85-89, ≥90 years). |
| Covid-19 vaccination status | *The Danish Vaccination Register*[3]. Vaccination status was defined by the number of received Covid-19 vaccines during the study period and status changed on the day of vaccination (5 levels: unvaccinated and having received |

| | |
|---|---|
| | 1, 2, 3, or 4 vaccine doses). We right-censored individuals at the date of receiving an Ad26.COV2-S Covid-19 vaccine as these were few in our study population (Ad26.COV2-S was not included in the standard Danish vaccination programme and recommended against for the studied age groups). For subgroup analysis, we categorized individuals' person-time according to whether being unvaccinated, primary course vaccinated (i.e., two vaccine doses), or booster vaccinated (i.e., first or second booster vaccine dose). Any fifth dose was considered as a right-censoring event as a fifth vaccine dose was not implemented in the general Danish rollout strategy of the Covid-19 vaccines during the study period. |
| Calendar time | Time was split every two weeks from study start (1 January 2021) to end of study period (10 December 2022) resulting in 51 distinct calendar time periods. |

ICD-10 denotes International Classification of Diseases System, version 10.

**Supplementary Table S8. Infectious disease hospitalization outcome definitions.**

| Outcome | Subcategory | ICD-10 codes |
|---|---|---|
| **Main outcome** | | |
| Any infection[a] | NA | NA |
| **Secondary outcomes** | | |
| Upper respiratory tract infection[b] | Abscessus peritonsillaris | J36 |
| | Ethmoiditis | J012 |
| | Infections in the ear | H65, H66, H67, H680, H70 |
| | Laryngitis | J370, A362, J04, J05 |
| | Nasopharyngitis | A361, J00 |
| | Other acute upper respiratory infections | A368, A369, J06 |
| | Pharyngitis | J02 |
| | Sinusitis | J32, J010, J011, J013, J014, J018, J019 |
| | Tonsillitis | J350, A360, J03 |
| Influenza | Influenza | J09, J10, J11 |
| Lower respiratory tract infection | Other acute lower respiratory infections | A37, A420, J20, J21, J22, J85, J86 |
| | Pneumonia | A481, A70, J12, J13, J14, J15, J16, J17, J18 |
| Gastrointestinal infection | NA | A00, A01, A020, A022, A028, A029, A421 |
| Skin infections | Cellulitis and abscess | H600, H601, L02, L03 |
| | Dermatophytosis and other superficial mycoses | B35, B36 |
| | Erysipelas | A46 |
| | Other local infections of skin and subcutaneous tissue | A363, H602, H603, H604, H608, H609, L00, L010, L011, L08, L303 |
| | Viral warts | B07 |
| Urinary tract infections | Cystitis | N300 |
| | Infections of the urinary system | N340, N341, N343 |
| | Pyelonephritis | N109 |
| Certain invasive bacterial infections | Infections of the circulatory system | I301, I33, I38, I398, I399, I400 |
| | Osteomyelitis | M462, M465, M860, M861, M862 |
| | Meningitis | A321, A390, A87, B003, B004, B010, B011, B020, B021, B050, B051, B060, B261, B262, G00, G01, G02, G03 |
| | Sepsis | A021, A327, A392, A40, A41, A499A |
| Other infections | Acute lymphadenitis | L04 |
| | Certain bacterial diseases | A2, A30, A31, A320, A328, A329, A33, A34, A35, A38, A391, A393, A394, A395, A398, |

| | |
|---|---|
| | A399, A422, A427, A428, A429, A43, A44, A480, A482, A483, A484, A488, A489, A490-A499 (not A499A), A50, A51, A52, A53, A54, A55, A56, A57, A748, A749 |
| Infections of the eye | A71, A72, A73, A740, H00, H010, H018, H019, H030, H031, H061, H100, H102, H103, H105, H106, H108 |
| Infections of the musculoskeletal system and connective tissue | M00, M01, M600, M863, M864, M865, M866 |
| Infections of the nervous system | G04, G05, G06, G07 |
| Hepatitis | B15, B16, B17, B18, B19 |
| Mycoses | B370, B371, B372, B373, B374, B375, B376, B378, B379, B38, B39, B4, B377 |
| Nephritis | N080 |
| Protozoal diseases helminthiases pediculosis acariasis and other infestations | A59, B5, B6, B7, B8 |
| Other types of other infections | A638, A639, A64 |
| Rickettsiosis | A75, A76, A77, A78, A79 |
| Spirochaetal disease | A65, A66, A67, A68, A69 |
| Tuberculosis | A15, A16, A17, A18, A19, K930 |
| Unspecified infections | B99 |
| Viral infections | A58, A60, A630, A80, A81, A82, A83, A84, A85, A86, A88, A89, A9, B000, B001, B002, B005, B006, B007, B008, B009, B012, B018, B019, B022, B023, B027, B028, B029, B03, B04, B052, B053, B054, B058, B059, B061, B068, B069, B08, B09, B20, B21, B22, B23, B24, B25, B260, B263, B268, B269, B27, B28, B29, B20, B31, B32, B338, B340, B341, B343, B344 |

ICD-10 denotes International Classification of Diseases System, version 10 and NA not applicable. Diagnoses were identified through use of the Danish National Patient Register.[4] Incident cases of infectious disease hospitalization was defined as hospital contacts of ≥5 hours (considered as inpatient contacts) and included both primary and secondary diagnoses. The admission date served as the event date. We excluded individuals with a registered hospital contact (regardless of the duration of the contact) with the respective outcome under studied from 1 January 2018 to the follow-up period. [a]Any infectious disease hospitalization include all outcomes and outcome definitions below. [b] Tonsillitis (ICD-10 codes: J350, A360, J03) was also studied separately (see Table S5).

### Supplementary Figure S2. Schematic figure of the study design.

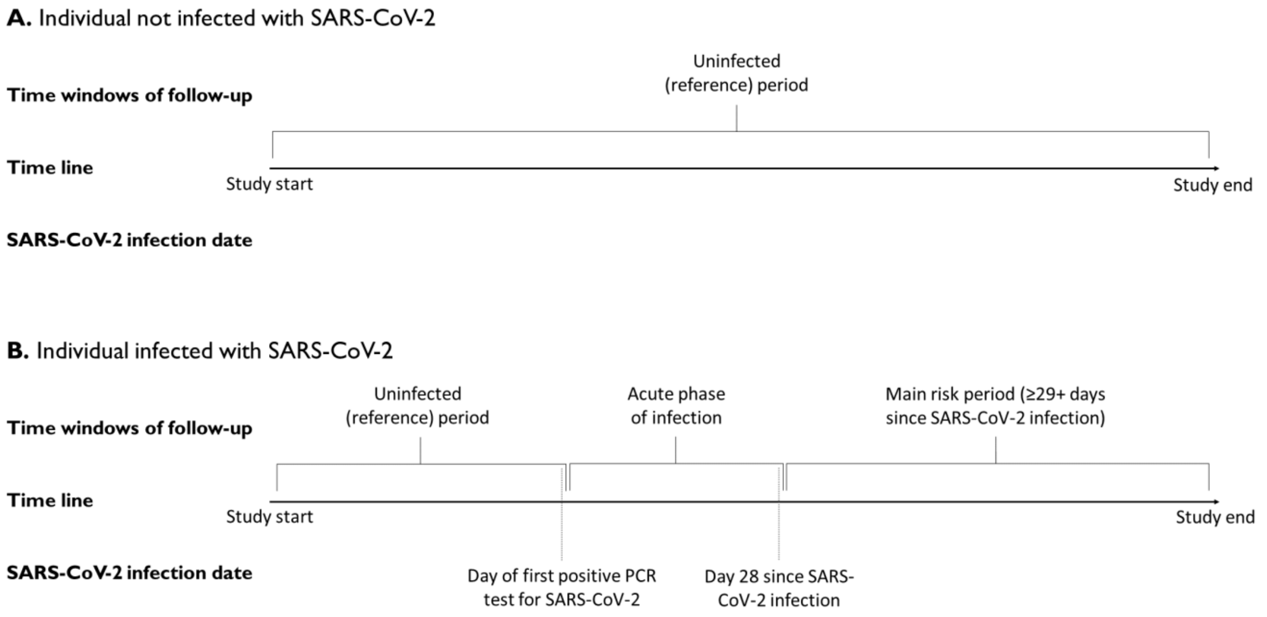


Start of follow-up was 1 January 2021 or turning age 50 years, whichever occurred last. Individuals were followed until first incident of the respective outcome event, second positive PCR test for SARS-CoV-2, emigration/disappearance, death, receiving an Ad26.COV2-S Covid-19 vaccine, receipt of a fifth vaccine dose, or 10 December 2022, whichever occurred first. Individuals could contribute with person-time during both the uninfected (reference) and main risk (29+ days following first SARS-CoV-2 infection) period as long as no outcome event had occurred prior to the respective follow-up period. Outcome rates during the main risk was compared with the uninfected reference period rates. In sensitivity analyses, we 1) deferred the main risk period to 90+ since the first SARS-CoV-2 infection date and included a pre-risk period of -7 days prior the day of the first SARS-CoV-2 infection date; this pre-risk period was omitted from the reference period.

**Supplementary Figure S3. Schematic figure of the analyses comparing SARS-CoV-2 infected and uninfected reference according to vaccination status at time of infection.**

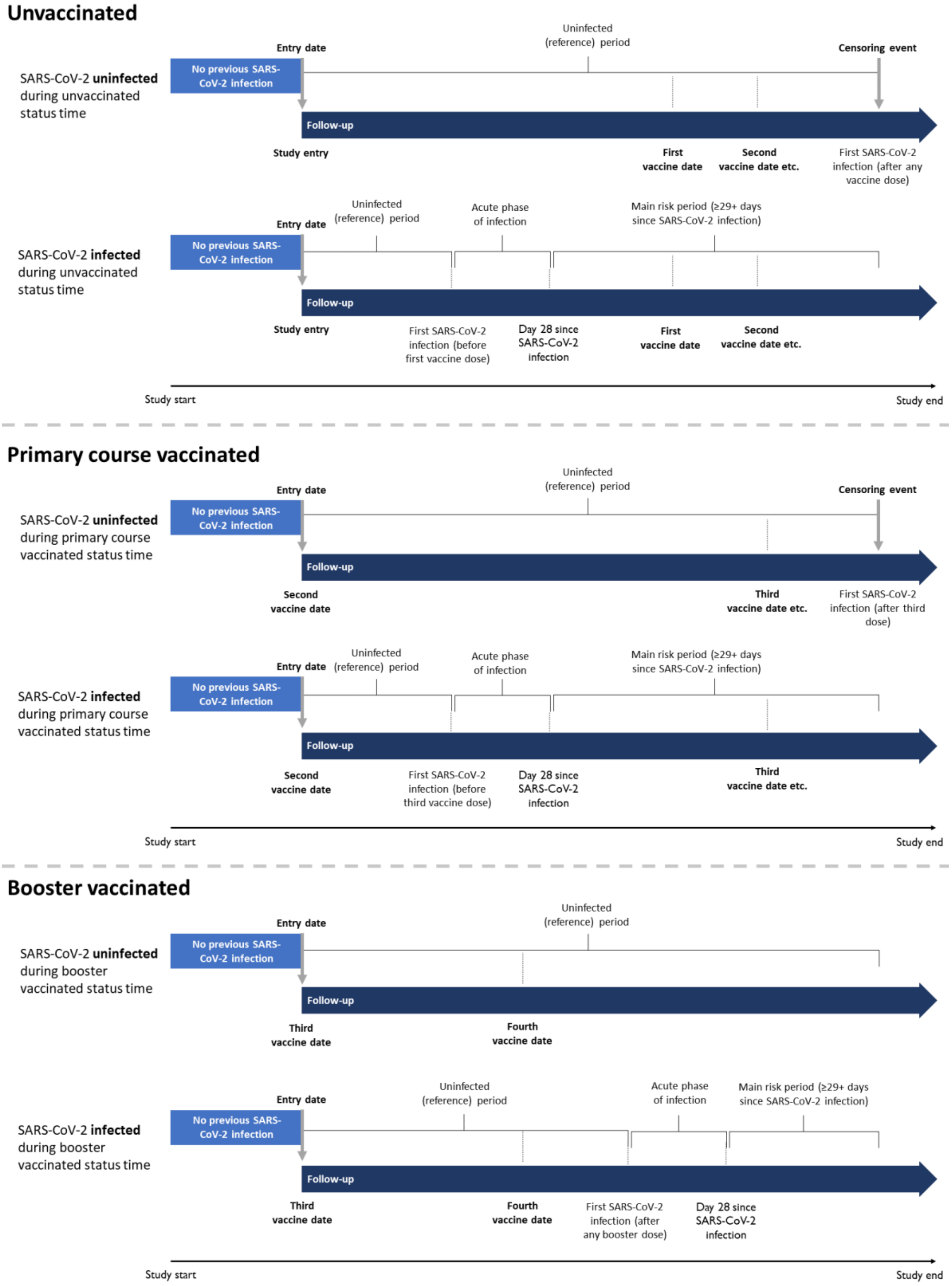


Each Covid-19 vaccination status at time of SARS-CoV-2 infection was analyzed separately (given the potential overlap of person-time across the three reference periods). Individuals were required to have no previous SARS-CoV-2 infection prior to the vaccination status-time analyzed and were followed up in a similar fashion as the main study design including the same censoring events. In addition, a first SARS-CoV-2 infection after any dose was treated as a censoring event for the unvaccinated comparison and after third dose for the primary course vaccinated comparison.
